# Mediterranean Dietary Approaches to Stop Hypertension Intervention for Neurodegenerative Delay Diet is Associated with Reduced Inflammatory Bowel Disease Related Surgery Risk: A Prospective Cohort Study

**DOI:** 10.64898/2026.05.28.26354274

**Authors:** Yuhao Sun, Zhuoyuan Jiang, Lintao Dan, Yuxin Qian, Judith Wellens, Jialu Yao, Xue Li, Xiaoyan Wang, Fernando Magro, Yang Chen, Jie Chen

**Author notes:** Correspondence: Jie Chen, PhD, Department of Gastroenterology, The Third Xiangya Hospital, Central South University, Changsha, China. Xiangya School of Public Health, Central South University, Changsha, Hunan, China. Postdoctoral Station of Clinical Medicine, the Third Xiangya Hospital, Central South University, Changsha, China., 4192, Yang Chen, MM, Department of Gastroenterology, The Third Xiangya Hospital, Central South University, Changsha, China. Nursing Department, The Third Xiangya Hospital, Central South University, Changsha, Hunan, China. Yuhao Sun, Zhuoyuan Jiang, and Lintao Dan contributed equally as co-first authors.

## Abstract

**Objectives:** The Mediterranean-DASH Intervention for Neurodegenerative Delay (MIND) diet has been associated with the risk of IBD, but its impact on clinical outcomes is uncertain. This study evaluated the association between MIND diet adherence and the risk of IBD-related surgery in a prospective cohort.

**Methods:** This study included 2,288 participants with diagnosis of Crohn’s disease (CD, n=777) or ulcerative colitis (UC, n=1,511) who completed valid WebQ 24-hour dietary recall from the UK Biobank. Dietary adherence was derived from a 15-component score based on 24-hour dietary recalls. Associations with IBD-related surgery were evaluated using Cox proportional hazards models, with nonlinear trends and examined via restricted cubic splines. Effect modification was explored in pre-specified subgroups, and multiple sensitivity analyses were conducted to assess robustness.

**Results:** During 10.9 years of follow-up, 166 incident IBD-related surgery cases occurred. Higher MIND diet adherence was associated with reduced surgical risk. Compared with the lowest tertile of adherence, the highest tertile showed a 36% reduction in surgical risk in IBD (HR 0.64, 95% CI: 0.44-0.94, *P* = 0.024). Notably, this protective effect was pronounced in patients with CD, exhibiting a clear linear inverse association. In contrast, a reverse J-shaped association was observed in UC, with a steep initial decline in surgical risk followed by a plateau emerging at a MIND score of approximately 5, beyond which further adherence conferred minimal additional benefit. At the component level, higher vegetable consumption and lower intake of butter and fried foods were identified as independent protective factors against surgery. Stronger inverse associations were observed among patients with shorter disease duration and those with complicated disease behavior, including stricturing or penetrating phenotypes (all *P* _interaction_ < 0.05).

**Conclusion:** Greater MIND diet adherence is associated with reduced IBD-related surgery risk among patients with IBD and CD. These findings support the MIND diet as a feasible dietary strategy to improve IBD prognosis.

## 1 Introduction

Inflammatory bowel disease (IBD), encompassing Crohn’s disease (CD) and ulcerative colitis (UC), is a chronic, relapsing inflammatory disorder of the gastrointestinal tract. Although the introduction of biologic therapies over the past decade has markedly reduced surgical intervention rates in both CD and UC, ^1–7^ the cumulative burden of IBD-related surgery remains considerable. Recent data have shown that the 10-year surgical risks have been 9.6% for UC and 26.2% for CD since the 20th century.^1^

Diet has emerged as a key modifiable environmental factor in IBD management, with current research exploring both isolated dietary components and comprehensive dietary patterns encompassing multiple nutritional elements.^8^ Exclusive enteral nutrition (EEN), partial enteral nutrition (PEN), and the Crohn’s Disease Exclusion Diet (CDED) demonstrate efficacy for inducing remission in active disease,^9–13^ yet these highly restrictive dietary strategies often compromise poor long-term adherence and quality of life.^14–16^ Furthermore, evidence for dietary maintenance of remission remains limited.^17,18^ Consequently, many patients empirically adopt unsubstantiated dietary exclusions, increasing risks for malnutrition and avoidant/restrictive food intake disorder without proven benefit.^19^ These challenges underscore the imperative for sustainable, nutritionally adequate dietary strategies concurrently with anti-inflammatory benefits.

The Mediterranean-Dietary Approach to Stop Hypertension Intervention for Neurodegenerative Delay (MIND) diet, integrating key components of the Mediterranean and DASH diets, is characterized by high intake of plant-based foods (such as leafy greens, berries, whole grains) and restriction of saturated fats and processed foods.^20,21^ Although originally developed for neurodegenerative disease prevention, its anti-inflammatory property and potential to modulate gut microbiota composition align closely with IBD pathophysiology.^22^ While observational studies have linked the MIND diet to reduced systemic inflammation and lower risk of non-communicable diseases (including IBD),^22–28^ its potential impact on IBD prognosis, particularly surgical risk, has not been systematically explored.

To address this gap, we utilized the UK Biobank cohort to investigate the longitudinal relationship between MIND diet adherence and IBD-related surgery. We hypothesized that higher adherence would confer a protective effect against IBD-surgery, with distinct patterns potentially emerging across IBD subtypes. Such findings could provide evidence-based insights to refine dietary recommendations for the long-term management of IBD.

## 2 Methods

### 2.1 Study Design and Participants

This prospective cohort study utilized data from the UK Biobank, a large population-based cohort comprising over 500,000 participants aged 37-73 years recruited from 22 assessment centers across England, Wales, and Scotland between 2006 and 2010.^29^ The UK biobank’s comprehensive data included touch-screen questionnaires, face-to-face interviews, physical measurements, biological sample collection, and linkage to electronic health records. Ethical approval was obtained from the North West-Haydock Research Ethics Committee (REC reference: 21/NW/0157).

Individuals with IBD were identified from self-reported data, primary care records, and hospital inpatient records. Identification of IBD subtypes is based on the corresponding codes from the International Classification of Disease (ICD)-9 and ICD-10 systems (ICD-9: 555, 556; ICD-10: K50, K51; **Table S1**).^30^

We identified 210,962 participants completing at least one recall of web-based 24-h dietary assessment in the UK Biobank. After excluding participants with implausible energy intake (defined as outside the range of 800-4200 kcal/day for men and 600-3500 kcal/day for women, n = 7,390) or reporting non-typical dietary recalls (n = 17,584),^31,32^ 185,988 participants with valid dietary records remained. Baseline was defined as the date of the last completed 24-h dietary recall, and then we included 2,288 individuals with baseline IBD (777 CD, 1,511 UC) for primary analysis (**Figure 1**).

**Figure 1.**
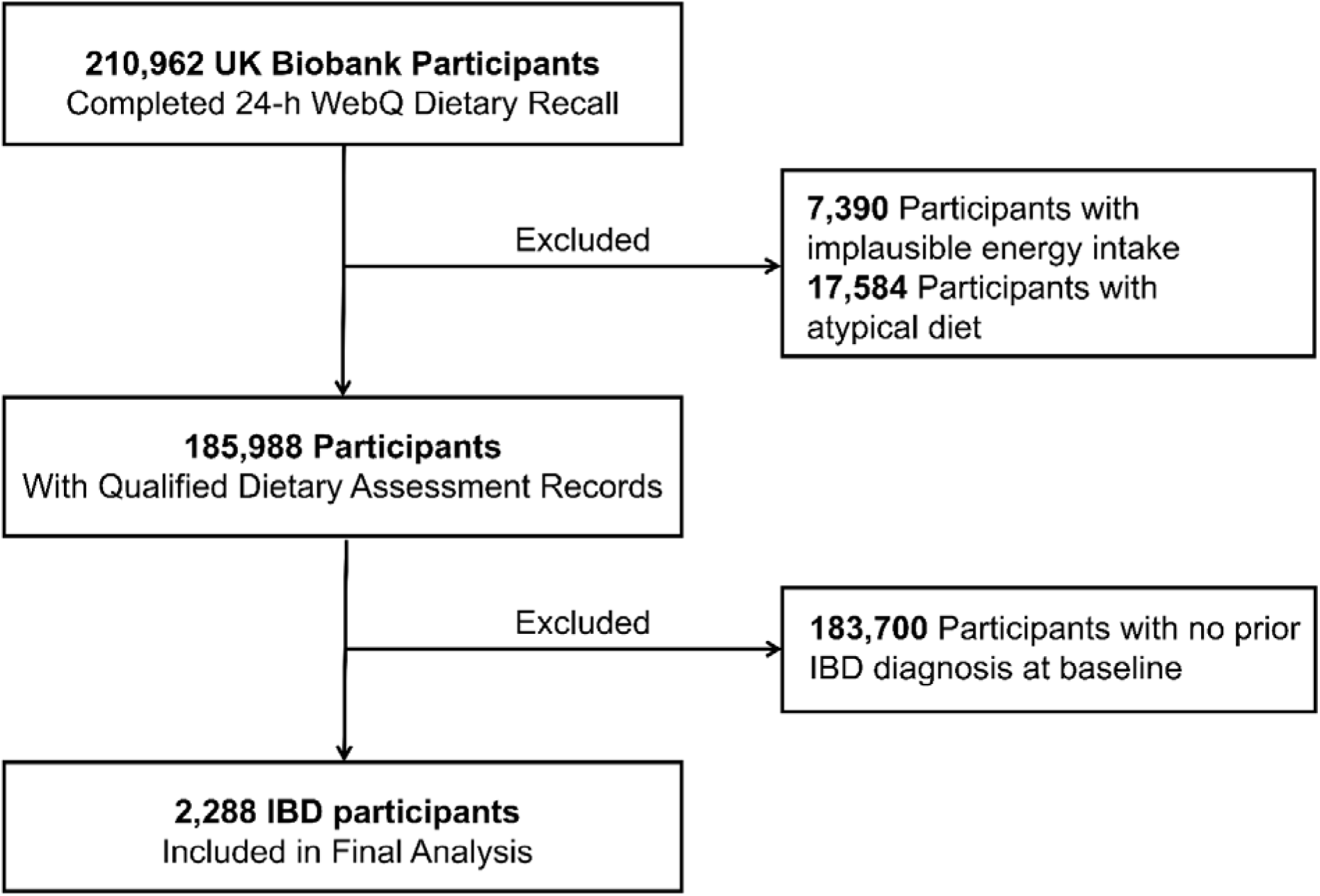
Participants inclusion.

### 2.2 Assessment of MIND Diet

Dietary intake was assessed using the validated Oxford Web-based 24-hour dietary recall questionnaire, including approximately 200 commonly consumed foods and beverages, which was administered on five occasions between 2009-2012.^33,34^ Dietary data derived from the validated 24-hour dietary recall questionnaires were calculated as averages to reduce potential measurement bias.

The MIND diet score was calculated based on 15 dietary components (10 beneficial, 5 detrimental) as previously described.^20–22^ Beneficial components included green leafy vegetables, other vegetables, berries, whole grains, non-fried fish, non-fried poultry, beans, nuts, olive oil (primary cooking oil), and wine. Detrimental components included red meat, butter, cheese, pastries/sweets, and fried foods. Each component was scored 0, 0.5, or 1 according to consumption frequency, except for olive oil, which was scored as 1 if reported as the primary cooking oil and 0 otherwise, similar to previous studies.^21,22^ Details are presented in **Table S2**. The MIND diet score ranged from 0 to 15, with higher scores reflecting greater adherence to the MIND diet.

### 2.3 Ascertainment of IBD-related surgery

IBD-related surgery was identified from hospital inpatient records using Office of Population Censuses and Surveys Classification of Interventions and Procedures (OPCS)-4 codes,^35^ including small bowel resection, colorectal resection, and proctological procedures. Details are presented in **Table S1**. The Audit Commission review of 2009 to 2010 concluded procedural coding OPCS-4 overall accuracy of 90% and diagnostic coding ICD-10 overall accuracy of 89%.^36^

### 2.4 Covariates

Potential variables were selected based on established literature and clinical relevance to MIND diet adherence and IBD prognosis.^37–39^ Demographic factors included age at recruitment (continuous), sex (male/female), ethnicity (White/others), and education level (college/below college). Socioeconomic status was assessed using the Townsend deprivation index (TDI), a validated measure of social deprivation in the UK population.^40^

Lifestyle factors comprised smoking status (never/previous/current), physical activity level (categorized as low, moderate, or high),^41^ total daily fiber intake (g/day), and total daily energy intake (kcal/day). Physical activity was collected from 24-hour dietary recall questionnaires and estimated by the Metabolic Equivalent of Task (MET). Total daily fiber intake and energy intake were also derived from 24-hour dietary recall questionnaires, with fiber intake assessed and summarized according to NHANES guidelines.^42^

Clinical characteristics included body mass index (BMI, kg/m²), Charlson comorbidity index (CCI, a weighted score of 17 chronic conditions),^43^ and IBD-specific factors such as disease subtype (CD/UC), disease duration (years since diagnosis), baseline history of IBD-related surgery (with and without), INFLA score (a composite marker of systemic inflammation derived from white blood cell count, platelet count, neutrophil-to-lymphocyte ratio, and C-reactive protein),^44,45^ use of IBD-related medications (aminosalicylates, corticosteroids, immunomodulators),^46^ as well as the presence of obstruction or stricture, and fistula or perforation.

Missing data for key covariates were minimal (<1%), and primary analyses employed single imputation (median for continuous variables, mode for categorical variables), with sensitivity analyses using multiple imputation to assess robustness of the results. More detailed information about covariables was shown in **Table S3**, and the condition of covariable missing was shown in **Table S4**.

### 2.5 Statistical analysis

Baseline characteristics were summarized by MIND diet score tertiles, with continuous variables reported as means (standard deviation, SD) and categorical variables as numbers (percentages). Group differences were assessed using nonparametric tests (Kruskal-Wallis) for continuous variables and chi-square tests for categorical variables. Follow-up time was calculated from baseline (defined as the completion date of the last 24-hour dietary recall) to the earliest of first recorded IBD-related surgery, death, loss to follow-up, or study end (England: 31 October 2022; Scotland: 31 August 2022; Wales: 31 May 2022). Participants with a history of IBD-related surgery prior to baseline were remained in the analysis, with baseline surgery status further incorporated as a stratification factor in subgroup analyses.

Cox proportional hazards models were used to estimate hazard ratios (HRs) with 95% confidence intervals (CIs), examining MIND diet scores both as tertiles and continuous variables (per 3-point increment).^47^ Two models were employed: Model 1 adjusted for age and sex, while Model 2 further adjusted for ethnicity, education level, BMI, TDI, smoking status, physical activity, and total energy intake. The proportional hazards assumption was verified using Schoenfeld residuals (all *P* > 0.49). Potential nonlinear associations were examined using restricted cubic spline models with knots at the 10th, 50th, and 90th percentiles. Kaplan-Meier curves were generated to visualize cumulative incidence across diet tertiles.

Secondary analyses were conducted further to assess the robustness and specificity of the primary findings. First, we performed component-level analyses to examine the independent associations between each MIND diet component and the risk of IBD-related surgery. We also evaluated associations stratified by anatomical sites of surgery, categorized as small-bowel resection, colorectal resection, and proctological procedures. In addition, analyses were stratified by hospital admission source, distinguishing elective from emergency surgeries.

Subgroup analyses were subsequently performed to evaluate potential effect modification. Stratification variables included (1) demographic factors: age (<60 vs. ≥60 years), sex (female vs. male), and BMI (<30 vs. ≥30 kg/m²); and (2) IBD-specific factors: INFLA score (≤0 vs. >0), disease duration (≤15 vs. >15 years), baseline history of IBD-related surgery (yes vs. no), IBD-related medication use (yes vs. no), presence of obstruction or stricture (yes vs. no), and presence of fistula or perforation (yes vs. no). Interaction effects were assessed on both the additive and multiplicative scales. Multiplicative interaction was evaluated by testing the significance of product terms (exposure × subgroup) entered into the multivariable regression models. Additive interaction was quantified by calculating the relative excess risk due to interaction (RERI), attributable proportion (AP), and synergy index (SI), with variance estimation derived using the delta method.

To assess the robustness of the findings, a series of sensitivity analyses were conducted, including: (1) reclassifying the MIND diet score into quartiles; (2) excluding events within the first year of follow-up; (3) additionally adjusting for the CCI; (4) additionally adjusting for total daily fiber intake; (5) modifying the MIND diet score after excluding the wine component; (6) handling missing data using multiple imputation; (7) restricting the analysis to participants with stricter IBD diagnostic criteria (at least 2 medical records);^48^ and (8) accounting for competing risk of mortality using the Fine and Gray model.^49^

All analyses were performed using R software (version 4.2.1), and a two-sided *P* value <0.05 was deemed significant.

## 3 Results

Of the total 2,288 participants, we documented 166 cases of IBD-related surgery (69 in CD and 97 in UC) over a median follow-up of 10.9 years. Baseline characteristics for overall IBD patients (**Table 1**), for CD patients (**Table S5**), and for UC patients (**Table S6**) are presented. Among these participants, 1,170 (51.1%) were female, and the average age at recruitment was 56.9 years. At baseline, the mean MIND diet score was 5.8. Individuals with higher MIND diet scores were more likely to be female, have a lower BMI, possess a higher educational level, be non-smokers, engage in more physical activity, and have lower energy intake (all *P* < 0.001). Differences in baseline characteristics were also observed between participants with and without dietary data (*P* < 0.05; **Table S7**), and these variables were included as covariates in the multivariable models.

**Table 1.** Baseline characteristics stratified by three levels of MIND diet score in participants with inflammatory bowel disease.

| Characteristic | Overall (n=2288) | MIND diet score in tertiles |  |  |
| --- | --- | --- | --- | --- |
|  |  | Tertile 1 (n=963) | Tertile 2 (n=608) | Tertile 3 (n=717) |
| MIND diet score, mean (SD) | 5.8 (2.0) | 3.9 (1.0) | 6.0 (0.4) | 8.1 (1.0) |
| Age, mean (SD) | 56.9 (7.9) | 56.7 (8.1) | 57.0 (7.9) | 57.1 (7.5) |
| Sex, n (%) |  |  |  |  |
| Female | 1170 (51.1) | 402 (41.7) | 316 (52.0) | 452 (63.0) |
| Male | 1118 (48.9) | 561 (58.3) | 292 (48.0) | 265 (37.0) |
| Townsend deprivation index, mean (SD) | -1.6 (2.8) | -1.5 (2.9) | -1.6 (2.7) | -1.6 (2.9) |
| Body mass index, kg/m<sup>2</sup>, mean (SD) | 26.8 (4.6) | 27.2 (4.6) | 26.9 (4.8) | 26.2 (4.4) |
| Education, n (%) |  |  |  |  |
| College | 852 (37.2) | 302 (31.4) | 227 (37.3) | 323 (45.0) |
| Below college | 1436 (62.8) | 661 (68.6) | 381 (62.7) | 394 (55.0) |
| Ethnicity, n (%) |  |  |  |  |
| White | 79 (3.5) | 27 (2.8) | 27 (4.4) | 25 (3.5) |
| Others | 2209 (96.5) | 936 (97.2) | 581 (95.6) | 692 (96.5) |
| Smoking status, n (%) |  |  |  |  |
| Never | 1113 (48.6) | 344 (46.4) | 324 (50.5) | 445 (49.2) |
| Previous | 1020 (44.6) | 326 (44.0) | 272 (42.4) | 422 (46.6) |
| Current | 155 (6.8) | 71 (9.6) | 46 (7.2) | 38 (4.2) |
| Physical activity, n (%) |  |  |  |  |
| Low | 762 (33.3) | 374 (38.8) | 200 (32.9) | 188 (26.2) |
| Moderate | 740 (32.3) | 278 (28.9) | 207 (34.0) | 255 (35.6) |
| High | 786 (34.4) | 311 (32.3) | 201 (33.1) | 274 (38.2) |
| Total energy, KJ/d, mean (SD) | 8688.4 (2071.0) | 9009.6 (2118.1) | 8813.2 (2124.2) | 8336.9 (1939.0) |
| IBD-specific characteristic |  |  |  |  |
| INFLA score, mean (SD) | 1.7 (6.2) | 2.2 (6.0) | 1.9 (6.2) | 1.0 (6.3) |
| Duration of IBD, years, mean (SD) | 18.5 (13.4) | 18.9 (13.3) | 17.9 (13.4) | 18.6 (13.5) |
| Baseline IBD-related surgery history, n (%) | 344 (15.0) | 176 (18.3) | 91 (15.0) | 77 (10.7) |
| IBD-related medication, n (%) | 943 (41.2) | 423 (43.9) | 232 (38.2) | 288 (40.2) |
| With obstruction or stricture, n (%) | 1146 (50.1) | 481 (49.9) | 306 (50.3) | 359 (50.1) |
| With fistula or perforation, n (%) | 1024 (44.8) | 421 (43.7) | 278 (45.7) | 325 (45.3) |
| IBD subtype, n (%) |  |  |  |  |
| Crohn's disease | 777 (34.0) | 274 (37.0) | 219 (34.1) | 284 (31.4) |
| Ulcerative colitis | 1511 (66.0) | 467 (63.0) | 423 (65.9) | 621 (68.6) |

MIND, Mediterranean-Dietary Approaches to Stop Hypertension Intervention for Neurodegenerative Delay; SD, standard deviation; IBD, inflammatory bowel disease.

### 3.1 Primary analysis

Higher adherence to the MIND diet was inversely associated with risk of IBD-related surgery in IBD, CD, and UC (**Figure 2**). RCS analysis revealed distinct dose-response relationships between the MIND diet score and the risk of surgery. A linear inverse association was observed for CD (*P*-nonlinear = 0.430), indicating a progressive reduction in surgical risk with increasing MIND diet adherence. IBD and UC showed nonlinear associations (both *P*-nonlinear < 0.01; **Figure 3**), with reverse J-shaped curves. The surgical risk initially declined steeply with higher MIND scores before reaching a plateau effect among IBD and UC patients, suggesting a threshold of maximal protection beyond which comparable risk reduction is sustained with further dietary improvements. Specifically, the protective effect against surgery in IBD and UC peaked at a MIND score of 5.2. When stratifying by adherence tertiles, the highest MIND diet adherence was associated with a significantly lower risk of IBD-related surgery among individuals with IBD (HR = 0.64, 95% CI: 0.44-0.94, *P* = 0.024) and CD (HR = 0.44, 95% CI: 0.23-0.83, *P* = 0.012). For UC, we observed an inverse association in Model 1(HR _per 3-point increment_ = 0.72, 95% CI: 0.52-0.98, *P* = 0.035), but the association did not remain statistically significant in the full adjusted model (HR _per 3-point increment_ = 0.75, 95% CI: 0.54-1.03, *P* = 0.076; **Table 2**).

**Figure 2.**
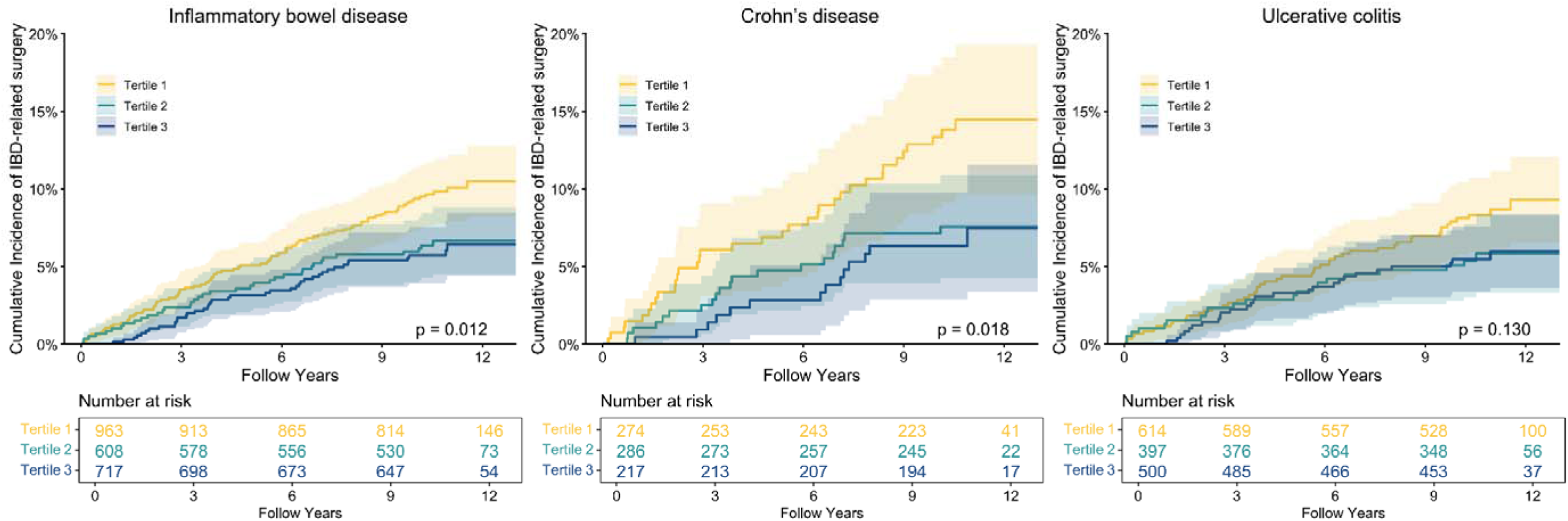
Kaplan-Meier curves for IBD-related surgery, stratified by tertiles of MIND diet score in patients with IBD, CD, and UC. MIND, Mediterranean-Dietary Approaches to Stop Hypertension Intervention for Neurodegenerative Delay; IBD, inflammatory bowel disease; CD, Crohn’s disease; UC, ulcerative colitis; HR, hazard ratios; CI, confidence interval.

**Figure 3.**
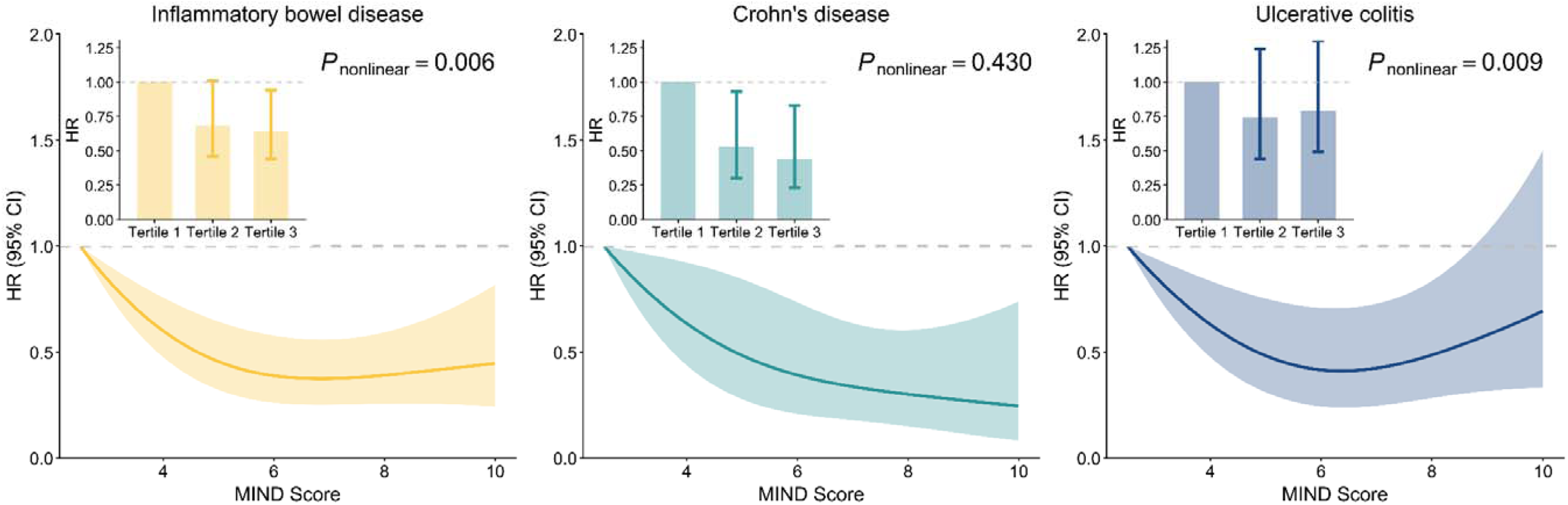
Dose-response relationships between MIND diet score and risk of IBD-related surgery in IBD, CD, and UC. The associations were examined using fully adjusted model with restricted curve splines. MIND, Mediterranean-Dietary Approaches to Stop Hypertension Intervention for Neurodegenerative Delay; IBD, inflammatory bowel disease; CD, Crohn’s disease; UC, ulcerative colitis; HR, hazard ratios; CI, confidence interval.

**Table 2.** Multivariable-adjusted hazard ratios and 95% confidence intervals for incident IBD-related surgery according to tertiles of MIND diet score.

| **MIND diet score** | **Cases/person-years** | **Model 1<sup>a</sup>**<br>**HR (95% CI)** | **P-value** | **Model 2<sup>b</sup>**<br>**HR (95% CI)** | **P-value** |
| --- | --- | --- | --- | --- | --- |
| **Inflammatory bowel disease** |  |  |  |  |  |
| Per 3-point increment |  | 0.63 (0.49, 0.80) | **<0.001** | 0.63 (0.49, 0.81) | **<0.001** |
| Tertile 1 | 88/9764 | 1 (Reference) |  | 1 (Reference) |  |
| Tertile 2 | 37/6170 | 0.67 (0.46, 0.99) | **0.042** | 0.68 (0.46, 1.01) | 0.054 |
| Tertile 3 | 41/7415 | 0.62 (0.43, 0.91) | **0.014** | 0.64 (0.44, 0.94) | **0.024** |
| P-trend |  |  | **0.009** |  | **0.016** |
| **Crohn's disease** |  |  |  |  |  |
| Per 3-point increment |  | 0.55 (0.37, 0.80) | **0.002** | 0.52 (0.35, 0.78) | **0.001** |
| Tertile 1 | 35/2733 | 1 (Reference) |  | 1 (Reference) |  |
| Tertile 2 | 20/2851 | 0.53 (0.31, 0.92) | **0.024** | 0.53 (0.30, 0.93) | **0.026** |
| Tertile 3 | 14/2247 | 0.46 (0.25, 0.86) | **0.015** | 0.44 (0.23, 0.83) | **0.012** |
| P-trend |  |  | **0.014** |  | **0.012** |
| **Ulcerative colitis** |  |  |  |  |  |
| Per 3-point increment |  | 0.72 (0.52, 0.98) | **0.035** | 0.75 (0.54, 1.03) | 0.076 |
| Tertile 1 | 49/6300 | 1 (Reference) |  | 1 (Reference) |  |
| Tertile 2 | 21/4051 | 0.70 (0.42, 1.18) | 0.182 | 0.74 (0.44, 1.24) | 0.253 |
| Tertile 3 | 27/5168 | 0.74 (0.46, 1.19) | 0.216 | 0.79 (0.49, 1.30) | 0.360 |
| P-trend |  |  | 0.182 |  | 0.315 |

MIND, Mediterranean-Dietary Approaches to Stop Hypertension Intervention for Neurodegenerative Delay; HR, hazard ratios; CI, confidence interval.
a. Model 1 adjusted for age and sex.
b. Model 2 additionally adjusted for Model 1, ethnicity, education, BMI, TDI, smoking status, physical activity, and total energy.

### 3.2 Secondary analysis

Associations between individual dietary components and the risk of IBD-related surgery are shown in **Table S8**. Higher intake of other vegetables (HR = 0.62, 95% CI: 0.40-0.96, *P* = 0.031), as well as lower intake of butter (HR = 0.63, 95% CI: 0.41-0.96, *P* = 0.031) and fried foods (HR = 0.49, 95% CI: 0.34-0.70, *P* < 0.001), was associated with reduced IBD-related surgery risk among individuals with IBD. These protective associations were consistently observed in CD (all *P* < 0.001), whereas in UC the associations were restricted to low consumption of fried foods (*P* = 0.004).

Site-specific analyses revealed distinct patterns of association between MIND diet adherence and surgical risk across IBD subtypes (**Table S9**). In the overall IBD population, higher adherence to the MIND diet showed significant inverse associations with the risks of small bowel resection, colorectal resection, and proctological surgery (**Figure 4**). When comparing extreme tertiles, the MIND diet was associated with a 56% lower risk of proctological surgery (HR _tertile 3 vs tertile 1_ = 0.44, 95% CI: 0.21-0.92, *P* = 0.029) in IBD and a 62% reduction in colorectal resection risk among patients with CD (HR _tertile 3 vs tertile 1_ = 0.38, 95% CI: 0.15-0.95, *P* = 0.039). UC showed significant associations only for proctological surgery, where each 3-point increment in MIND diet score was associated with a lower risk (HR = 0.47, 95% CI: 0.24-0.92, *P* = 0.028).

**Figure 4.**
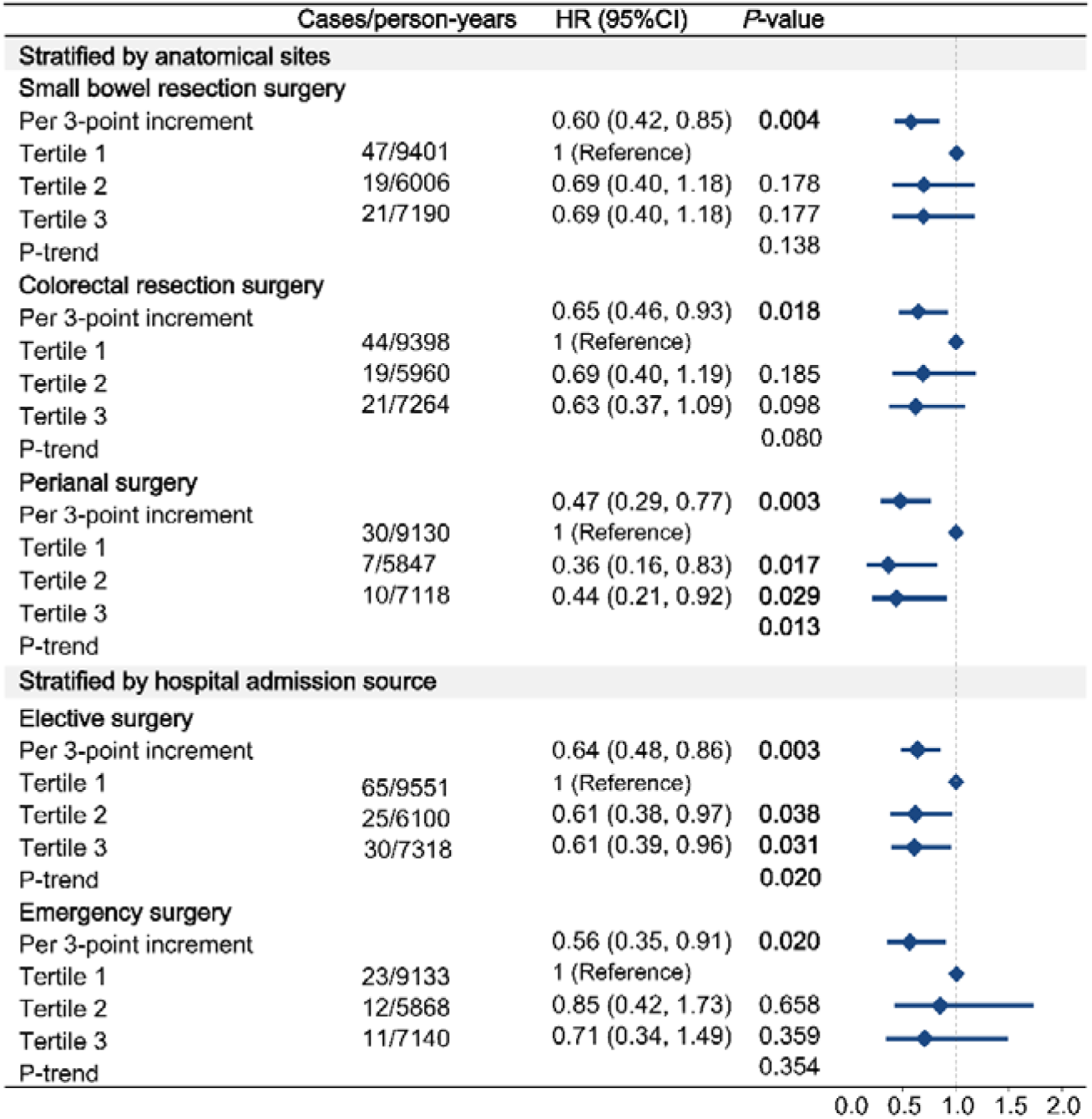
Forest plot of associations between the MIND diet and risk of IBD-related surgery in IBD, stratified by anatomical sites of surgery and hospital admission source.

Further analyses examined the association between the MIND diet and surgery risk stratified by admission source. Higher adherence to the MIND diet was associated with a reduced risk of both elective and emergency surgeries among patients with IBD and CD (*P* _per 3-point increment_ < 0.05), with no corresponding association observed in UC (**Table S10**). In tertile analyses, patients with IBD in the highest adherence had a 39% lower risk of elective surgery compared with those in the lowest tertile (HR = 0.61, 95% CI: 0.39-0.96, *P* = 0.031; **Figure 4**). For emergency surgery, a significant protection was observed only in CD, with the highest adherence tertile associated with a lower risk (HR = 0.26, 95% CI: 0.07-0.94, *P* = 0.041). Across all models, UC showed no significant associations with either elective or emergency surgery.

### 3.3 Subgroup analysis

Subgroup analyses stratified by demographic and clinical characteristics are presented in **Tables S11-13**. Effect modification was evaluated on both the multiplicative and additive scales. The multiplicative scale assesses heterogeneity in relative risks across subgroups, whereas the additive scale measures the excess absolute risk attributable to the combined exposure, thereby identifying synergistic or antagonistic joint effects. Among demographic factors, significant effect modification by sex was observed among patients with CD (*P* for multiplicative interaction = 0.035), with stronger inverse associations between MIND diet adherence and surgical risk in females. No evidence of interaction was observed for age or BMI (Both multiplicative and additive interaction *P* > 0.05).

Regarding clinical characteristics, disease duration modified the association on the multiplicative scale (*P* = 0.047), with stronger associations observed in patients with disease duration less than 15 years. Disease complications also demonstrated significant effect modification. On the multiplicative scale, the presence of complications (including stricture and perforation) altered the association between MIND diet adherence and surgical risk (all *P* for multiplicative interaction < 0.05), with greater surgical risk reduction observed in patients with complicated disease behavior. On the additive scale, significant interactions between MIND diet adherence and perforation were identified in both IBD (RERI = 1.08, 95% CI: 0.21-1.95, *P* = 0.008) and CD (RERI = 3.40, 95% CI: 0.52-6.29, *P* = 0.010; **Table S14**), indicating a greater benefit of the MIND diet in IBD patients with complicated disease behavior. No effect modification was observed for inflammation severity, baseline surgery history, or use of IBD-related medications.

### 3.4 Sensitivity analysis

The observed results remained consistent across most sensitivity analyses (**Tables S15-16**). Similar associations were observed when the MIND diet score was modeled in quartiles and when recalculated with the wine component excluded, confirming the robustness of the findings under alternative exposure definitions. Adjustment for additional covariates did not alter the results, including further control for the CCI and total fiber intake, accounting for the competing risk of mortality, and handling missing data using multiple imputation. The protective association persisted after excluding events occurring within the first year of follow-up and when limiting the analysis to participants meeting stricter IBD diagnostic criteria.

## 4 Discussion

In this large prospective cohort of patients with IBD, higher adherence to the MIND diet was consistently associated with a lower risk of IBD-related surgery, with a more pronounced protective effect in CD than in UC. Dose-response analyses further revealed subtype-specific patterns, with a linear association in CD and a reverse J-shaped relationship in overall IBD and UC, suggesting a plateau of benefit at moderate adherence levels in the latter. These findings extend prior evidence linking the MIND diet to reduced IBD incidence and highlight its potential role in improving long-term disease prognosis.

The distinct dose-response relationships between CD and UC likely reflect their fundamental pathophysiological differences. CD is characterized by progressive structural damage, which may be more amenable to cumulative modification through sustained dietary improvement. In contrast, UC involves superficial, continuous mucosal inflammation, in which dietary interventions are more likely to reach a plateau in their effects. Similar patterns have been observed for prior study of dietary total antioxidant capacity, which has demonstrated stronger associations with CD than with UC.^50^

Notably, the protective association of the MIND diet was stronger among patients with complicated disease behavior, including stricturing or penetrating phenotypes, and showed significant additive and multiplicative interactions with perforation in overall IBD and CD. This suggests that dietary modulation may be particularly relevant in patients with aggressive disease phenotypes and higher inflammatory burden.^51^ In this high-risk population, the MIND diet may serve as a complementary strategy to pharmacological therapy by attenuating inflammation and potentially delaying disease progression, thereby reducing the need for surgical intervention.

From a clinical perspective, the MIND diet offers a pragmatic and sustainable approach to dietary management in IBD. Although restrictive interventions such as EEN and CDED are effective for inducing remission, their long-term applicability is limited by poor adherence and uncertain maintenance efficacy.^19^ In contrast, the MIND diet, grounded in the Mediterranean dietary framework, emphasizes increased consumption of anti-inflammatory foods while avoiding extreme dietary restriction. This balance enhances feasibility in real-world settings and aligns with emerging evidence that Mediterranean dietary patterns, particularly when combined with healthy lifestyle behaviors, are associated with reduced relapse risk and improved clinical outcomes in IBD.^17,18,52,53^

Among individual dietary components, higher vegetable intake and lower consumption of butter and fried foods were independently associated with reduced surgical risk. These findings reinforce the central role of plant-based, fiber-rich foods in modulating intestinal inflammation and are consistent with our previous work linking healthy plant-based dietary patterns to improved IBD surgical outcomes (HR = 0.50, 95% CI: 0.30-0.83, *P*-trend < 0.001).^54^ Higher fiber intake from vegetable sources was also associated with reduced IBD-related surgery risk, with a 23% lower risk observed in the highest intake group (HR = 0.77, 95% CI: 0.60-0.98, *P*-trend = 0.007).^35^ Consistently, prospective trials have shown that adherence to anti-inflammatory dietary patterns can support maintenance of remission in IBD.^55^ Conversely, butter and fried foods represent key elements of pro-inflammatory dietary patterns and have been associated with increased disease activity and symptom burden.^56^

Interestingly, these components differ from those implicated in IBD incidence,^22^ suggesting that dietary factors influencing disease progression and surgical outcomes may operate through pathways distinct from those implicated in disease initiation.^57^ Wine, which has been independently associated with lower IBD incidence, did not show a protective association with surgical outcomes in this cohort. Sensitivity analyses excluding wine from the MIND score produced similar results, suggesting that this component did not drive the observed benefit of the MIND diet.

To our knowledge, this is the first study to explore the association between the MIND diet and IBD-related surgery, and our research is based on a large-scale prospective cohort with over 10 years of follow-up. Despite these strengths, the study has several limitations. First, as an observational study, it cannot establish causality or exclude the possibility of residual confounding. To address this, we excluded incident cases in the first year of follow-up and used multivariate adjustment to avoid reverse causality. Second, the criteria for wine consumption and score in the MIND diet vary across studies, so we excluded the wine score and modified the MIND score to a 0-14 range in sensitivity analysis to minimize the impact of such methodological inconsistencies. Third, IBD patients with malnutrition or irregular diets differ from healthy individuals in energy intake and food patterns. Thus, we excluded the participants with extreme energy intake based on all IBD cases in the UK Biobank. Fourth, predominantly White, middle-aged demographic composition limits the generalizability of the findings. Further research in ethnically diverse populations is thus warranted.

In conclusion, higher adherence to the MIND diet was associated with a lower risk of IBD-related surgery, particularly among patients with CD and those with complicated disease behavior. Our findings suggest that the MIND diet represents a feasible and effective dietary strategy for reducing surgical risk and improving clinical outcomes in the IBD population, and it should be evaluated in future randomized controlled trials.

## Author contributions

All authors have read and agreed to the published version of the manuscript. Yuhao Sun: conceptualization (equal); methodology (equal); visualization (supporting); writing original draft (equal); writing review and editing (leading); Lintao Dan: conceptualization (equal); methodology (supporting); visualization (equal); writing original draft (equal); writing review and editing (supporting); Zhuoyuan Jiang: conceptualization (equal); methodology (supporting); visualization (leading); writing original draft (leading); writing review and editing (equal); Yuxin Qian: methodology (equal); writing review and editing (supporting); Judith Wellens: methodology (supporting); writing review and editing (supporting); Jialu Yao: methodology (supporting); writing review and editing (supporting); Xue Li: methodology (supporting); writing review and editing (supporting); Xiaoyan Wang: methodology (supporting); writing review and editing (supporting); project administration (leading); Fernando Magro: methodology (supporting); writing review and editing (supporting); Yang Chen: methodology (supporting); writing review and editing (supporting); Jie Chen: conceptualization (leading); data curation (leading); formal analysis (leading); writing review and editing (equal).

## Ethical approval and consent

Ethical approval for the UK Biobank was obtained from the North West-Haydock Research Ethics Committee (REC reference: 21/ NW/0157), and all participants provided written informed consent prior to inclusion in the study.

## Data Availability

The datasets analyzed in this study are publicly available through the UK Biobank (https://www.ukbiobank.ac.uk).

## Acknowledgements

This work was conducted using the UK Biobank Resource under application number 232231. We want to thank all UK Biobank participants and the management team for their participation and assistance. All authors approved the final version of the article, including the authorship list.

