## Supplement for "Mediterranean Dietary Approaches to Stop Hypertension Intervention for Neurodegenerative Delay Diet is Associated with Reduced Inflammatory Bowel Disease Related Surgery Risk: A Prospective Cohort Study"

Table S1. Diagnostic codes of inflammatory bowel disease and IBD-related surgery.

|  | **Diagnostic Codes** | |
| --- | --- | --- |
| Disease | ICD-9 | ICD-10 |
| Crohn’s Disease | 555 | K50 |
| Ulcerative colitis | 556 | K51 |
| IBD-related surgery | OPCS4 | |
| Small bowel resection | G32-33, G343, G58-62, G632, G638, G639, G671, G672, G678, G679, G69-76 (except for G701, G733, G734, G764), G78-79 (except for G781), G801, G808, G809, G82 | |
| Large bowel and colorectal resection | H06, H07 | |
| Colorectal resection | H04, H05, H08-H12, H29, H308, H309, H31, H33, H469, H471, H478, H623, H624, H66, J011 | |

IBD, inflammatory bowel disease; ICD, International Classification of Diseases; OPCS, Office of Population Censuses and Surveys Classification of Interventions and Procedures.

### Table S2. MIND diet component servings and scoring.

| **MIND diet component** | **Example food items in the Oxford WebQ** | **Scoring** | | |
| --- | --- | --- | --- | --- |
|  |  | **0** | **0.5** | **1** |
| Green leafy vegetables | Cabbage/kale, lettuce, spinach | ≤2 servings/wk | >2 to<6 servings/wk | ≥6 servings/wk |
| Other vegetables | green beans, broccoli, butternut squash, carrot, cauliflower, celery, cucumber, leek, mushroom, sweet pepper, sprouts, sweetcorn, sweet potato | <5 servings/wk | 5 to<7 servings/wk | ≥1 servings/day |
| Berries | berries | <1 serving/wk | 1 serving/wk | ≥2 servings/wk |
| Nuts | nuts, peanuts, seeds, peanut butter | <1 serving/m | 1/m to<5 servings/wk | ≥5 servings/wk |
| Olive oil | Participants used olive oil in cooking across all diet records that reported using fat/oil in cooking. | No primary oil |  | Primary oil used |
| Butter/margarine | butter/margarine on bread/crackers and potatoes | >2 tsp/day | 1–2 tsp/day | <1 tsp/day |
| Cheese | hard cheese, soft cheese, cheese spread, cottage cheese, mozzarella, goat’s cheese | ≥7 servings/wk | 1 to 6 servings/wk | <1 servings/wk |
| Whole grains | porridge, whole-wheat cereal/breads, whole meal pasta, brown rice | <1 serving/m | 1–2 servings/d | ≥3 servings/d |
| Fish (not fried) | tinned tuna, oily fish, white fish, prawns, lobster/crab, shellfish | <1 servings/d | 1 to 3 servings/m | ≥1 servings/wk |
| Beans | baked bean, pulses, broad bean, hummus, tofu | <1 serving/wk | 1–3 servings/wk | >3 servings/wk |
| Poultry (not fried) | poultry | <1 serving/wk | 1 serving/wk | ≥2 servings/wk |
| Red meat and products | sausage, beef, pork, lamb, bacon, ham | ≥7 servings/wk | 4–6 servings/wk | <4 servings/wk |
| Fast/fried foods | fried poultry, breaded fish, fried potatoes, crisp/chips | ≥4 servings/wk | 1–3 servings/wk | <1 serving/wk |
| Pastries and sweets | pastry, crumble, pancake, pudding, ice-cream, cake, doughnut, chocolate bar, biscuits, hot chocolate, fizzy drink, added sugars and preserves | ≥7 servings/wk | 5–6 servings/wk | <5 servings/wk |
| Wine | red wine, rose wine, white wine, fortified wine | >1 glass/day or never | 2-6 glass/wk | 1 glass/day |

MIND, Mediterranean-Dietary Approaches to Stop Hypertension Intervention for Neurodegenerative Delay.

### Table S3. Definitions of covariables in the analysis.

| **Covariables** | **Definitions** |
| --- | --- |
| Age at recruitment | Continuous variable.  Age at recruitment is based on the participant's date of birth and date of attendance at the initial assessment. Refer to the age of the participant on the date of attendance at the initial assessment center, truncated to a full year. |
| Sex | Categorical variable (male, female).  Derived from central registry at recruitment. |
| Ethnicity | Categorical variable (white, other).  Ascertained via the UK Biobank touchscreen questionnaire on ethnic background. The "white" category includes British, Irish, and other white backgrounds. |
| Education | Categorical variable (college, below college).  Derived from the UK Biobank touchscreen questionnaire on qualification possession. The "below college" category includes A levels/AS levels or equivalent, O levels/GCSEs or equivalent, CSEs or equivalent, NVQ, HND, HNC or equivalent, other professional qualifications (e.g., nursing, teaching), and no qualifications, and none of the above. |
| Townsend deprivation index | Continuous variable.  Townsend deprivation index (TDI) was conducted as a complex indicator of socioeconomic status based on the unemployment rate, the percentage of overcrowded households, the percentage of people without cars, and the percentage of people without houses for each area in the UK. TDI was derived from the preceding national census output areas at recruitment using the method mentioned online. The higher TDI reflected the greater socioeconomic deprivation one was suffering.^(1)^ |
| Body mass index (BMI) | Continuous variable.  Calculated as weight divided by height squared (kg/m²). Height and weight data were collected via the UK Biobank touchscreen questionnaire, and BMI was derived therefrom. |
| Smoking status | Categorical variable (never, previous, current).  Ascertained via the UK Biobank touchscreen questionnaire on current/past smoking status. |
| Physical activity | Categorical variable (low, moderate, high).  The type and duration of physical activity from questions in 24-hour dietary recall questionnaire. Total physical activity was estimated by weighting the reported minutes of each activity category by the corresponding MET (Metabolic Equivalent of Task) energy expenditure estimate assigned to each category of activity (Light physical activity for 2 METs/h, moderate physical activity for 4.0 METs/h and vigorous physical activity for 8.0 METs/h) ^(2)^, and further classified into three categories, such as low, moderate and high. |
| Total daily energy | Continuous variable, kcal/day.  Total energy from overall diet is estimated from the mean intake of the 24-hour dietary recall questionnaire. |
| Total daily fiber intake | Continuous variable, g/day  Fiber intake for each food item from 24-hour dietary recall questionnaire, was calculated as consumption amount multiplied by its fiber content (from a food composition database). These individual fiber intakes were then summed to yield the total dietary fiber intake. |
| Charlson comorbidity index | Continuous variable, ranging from 0 to 16.  Charlson Comorbidity Index (CCI), is an indicator reflecting participants' objective health status and comorbidities, developed by Charlson et al.^(3)^ It is constructed based on 17 comorbidities identified via ICD codes from hospital records, including myocardial infarction, congestive heart failure, peripheral vascular disease, cerebrovascular disease, dementia, chronic pulmonary disease, rheumatic disease, peptic ulcer disease, mild liver disease, diabetes (with or without chronic complications), hemiplegia or paraplegia, renal disease, any malignancy (including lymphoma and leukemia but excluding cutaneous malignant neoplasms), moderate or severe liver disease, metastatic solid tumors, and acquired immunodeficiency syndrome. Each comorbidity category is assigned a weight based on the adjusted risk of mortality or resource use. A higher CCI score indicates a greater likelihood of predicted negative outcomes. It is a well-established, highly cited tool for quantifying comorbidity burden and evaluating impacts on health outcomes in clinical research. |
| INFLA score | Continuous variable, ranging from -16 to +16.  INFLA score is calculated based on white blood cells and platelet count, neutrophil-to-lymphocyte ratio, and C-reactive protein. These biomarkers, which were derived from the biology sample collected at baseline, were measured via blood count processing on a Beckman Coulter LH750 and high-sensitivity immunoturbidimetric analysis on a Beckman Coulter AU5800. Each biomarker is scored from −4 to +4 based on deciles: deciles 1–4 are assigned −4 to −1, deciles 5–6 score 0, and deciles 7–10 are assigned +1 to +4. It was a composite marker of systemic inflammation. ^(4, 5)^ |
| Duration of IBD | Continuous variable, years  The duration of IBD was calculated from the date of first diagnosis until the baseline date. |
| Baseline IBD-related surgery history | Categorical variable (free of surgery, with surgery history).  Baseline IBD-related surgery was defined as having been performed before recruitment, including small bowel resection, small bowel and colorectal resection, colorectal/large bowel resection, surgery for perianal disease, and other surgery. Surgeries were identified through inpatient data documented in the operations and procedures code according to the Office of Population Censuses and Surveys Classification of Interventions and Procedures (OPCS). |
| IBD-related medication | Medication use was identified from UK Biobank GP prescription records, specifically field *1062*. Participants were classified as “with medication” if they had prescriptions for aminosalicylates (Mesalazine, Sulfasalazine, Olsalazine, Balsalazide), corticosteroids (Budesonide, Dexamethasone, Hydrocortisone, Methylprednisolone, Prednisolone, Prednisone), or immunomodulators (Azathioprine, Mercaptopurine, Methotrexate, Cyclosporine A, Tacrolimus). Those without such prescriptions were classified as “free of medication.” This variable reflects baseline pharmacological management of IBD. |
| Obstruction or stricture | Categorical variable (without obstruction or stricture, with obstruction or stricture).  Diagnosis was ascertained from UK Biobank Hospital Episode Statistics, self-reports, the Death registry, and GP data records. Relevant codes included ICD-10: K56.4, K56.5, K56.6, K56.7, K62.4; and ICD-9: 560.8, 560.9. Participants with any of these codes were classified as “with obstruction or stricture.” This variable indicates structural complications of obstruction and stricture. |
| Fistula or perforation | Categorical variable (without fistula or perforation, with fistula or perforation).  Diagnosis was ascertained from UK Biobank Hospital Episode Statistics, self-reports, the Death registry, and GP data records. Relevant codes included ICD-10: K63.1, K63.2, K60.4; and ICD-9: 566. Participants with any of these codes were classified as “with fistula or perforation.” This variable reflects severe IBD-related complications, such as intestinal perforation and fistula formation, which are associated with adverse outcomes. |

### Table S4. Missing number and rates of covariates.

| **Covariates** | **Inflammatory bowel disease** | | **Crohn’s disease** | | **Ulcerative colitis** | |
| --- | --- | --- | --- | --- | --- | --- |
|  | **Number** | **Rate (%)** | **Number** | **Rate (%)** | **Number** | **Rate (%)** |
| Age | 0 | 0 | 0 | 0 | 0 | 0 |
| Sex | 0 | 0 | 0 | 0 | 0 | 0 |
| Education | 13 | 0.57 | 6 | 0.77 | 7 | 0.46 |
| Ethnicity | 2 | 0.09 | 1 | 0.13 | 1 | 0.07 |
| Townsend deprivation index | 2 | 0.09 | 2 | 0.26 | 0 | 0 |
| BMI | 6 | 0.26 | 2 | 0.26 | 4 | 0.26 |
| Smoking status | 2 | 0.09 | 1 | 0.13 | 1 | 0.07 |
| Physical activity | 0 | 0 | 0 | 0 | 0 | 0 |
| Total energy | 0 | 0 | 0 | 0 | 0 | 0 |
| Fiber intake | 0 | 0 | 0 | 0 | 0 | 0 |
| Charlson comorbidity index | 0 | 0 | 0 | 0 | 0 | 0 |
| INFLA score | 180 | 7.87 | 63 | 8.11 | 117 | 7.74 |
| Duration of IBD | 1 | 0.04 | 0 | 0 | 1 | 0.07 |
| Baseline IBD-related surgery history | 0 | 0 | 0 | 0 | 0 | 0 |
| IBD related medicatio | 0 | 0 | 0 | 0 | 0 | 0 |
| With obstruction or stricture | 0 | 0 | 0 | 0 | 0 | 0 |
| With fistula or perforation | 0 | 0 | 0 | 0 | 0 | 0 |

### Table S5. Baseline characteristics stratified by three levels of MIND diet score in participants with Crohn’s disease.

| **Characteristic** |  | **MIND diet score in tertiles** | | |
| --- | --- | --- | --- | --- |
|  | **Overall (n=777)** | **Tertile 1 (n=274)** | **Tertile 2 (n=286)** | **Tertile 3 (n=217)** |
| MIND diet score, mean (SD) | 5.6 (1.9) | 3.6 (0.9) | 5.7 (0.6) | 8.0 (1,0) |
| Age, mean (SD) | 55.9 (8.0) | 55.2 (8.3) | 55.9 (8.2) | 56.7 (7.1) |
| Sex, n (%) |  |  |  |  |
| Female | 416 (53.5) | 128 (46.7) | 151 (52.8) | 137 (63.1) |
| Male | 361 (46.5) | 146 (53.3) | 135 (47.2) | 80 (36.9) |
| Townsend deprivation index, mean (SD) | -1.7 (2.8) | -1.4 (2.9) | -1.8 (2.7) | -1.9 (2.6) |
| Body mass index, kg/m^2^, mean (SD) | 26.4 (4.5) | 26.8 (4.7) | 26.6 (4.8) | 25.7 (3.8) |
| Education, n (%) |  |  |  |  |
| College | 275 (35.4) | 80 (29.2) | 99 (34.6) | 96 (44.2) |
| Below college | 502 (64.6) | 194 (70.8) | 187 (65.4) | 121 (55.8) |
| Ethnicity, n (%) |  |  |  |  |
| White | 30 (3.9) | 7 (2.6) | 14 (4.9) | 9 (4.1) |
| Others | 747 (96.1) | 267 (97.4) | 272 (95.1) | 208 (95.9) |
| Smoking status, n (%) |  |  |  |  |
| Never | 359 (46.2) | 109 (39.8) | 144 (50.3) | 106 (48.8) |
| Previous | 345 (44.4) | 131 (47.8) | 116 (40.6) | 98 (45.2) |
| Current | 73 (9.4) | 34 (12.4) | 26 (9.1) | 13 (6.0) |
| Physical activity, n (%) |  |  |  |  |
| Low | 249 (32.0) | 101 (36.9) | 93 (32.5) | 55 (25.3) |
| Moderate | 270 (34.7) | 75 (27.4) | 114 (39.9) | 81 (37.3) |
| High | 258 (33.2) | 98 (35.8) | 79 (27.6) | 81 (37.3) |
| Total energy, KJ/d, mean (SD) | 8732.7 (2124.6) | 8922.3 (2159.9) | 8779.5 (2113.9) | 8431.9 (2069.9) |
| IBD-specific characteristic |  |  |  |  |
| INFLA score, mean (SD) | 2.7 (6.2) | 3.2 (6.0) | 3.2 (6.4) | 1.5 (6.1) |
| Duration of IBD, years, mean (SD) | 19.1 (13.4) | 19.0 (13.0) | 19.3 (13.7) | 19.0 (13.4) |
| Baseline IBD-related surgery history, n (%) | 174 (22.4) | 72 (26.3) | 65 (22.7) | 37 (17.1) |
| IBD related medication, n (%) | 331 (42.6) | 132 (48.2) | 112 (39.2) | 87 (40.1) |
| With obstruction or stricture, n (%) | 283 (36.4) | 102 (37.2) | 111 (38.8) | 70 (32.3) |
| With fistula or perforation, n (%) | 201 (25.9) | 74 (27.0) | 82 (28.7) | 45 (20.7) |

MIND, Mediterranean-Dietary Approaches to Stop Hypertension Intervention for Neurodegenerative Delay; SD, standard deviation; IBD, inflammatory bowel disease.

### Table S6. Baseline characteristics stratified by three levels of MIND diet score in participants with ulcerative colitis.

| **Characteristic** |  | **MIND diet score in tertiles** | | |
| --- | --- | --- | --- | --- |
|  | **Overall (n=1511)** | **Tertile 1 (n=614)** | **Tertile 2 (n=397)** | **Tertile 3 (n=500)** |
| MIND diet score, mean (SD) | 5.9 (2.0) | 3.9 (1.0) | 6.0 (0.4) | 8.1 (1.0) |
| Age, mean (SD) | 57.4 (7.78) | 57.4 (7.9) | 57.6 (7.7) | 57.3 (7.6) |
| Sex, n (%) |  |  |  |  |
| Female | 754 (49.9) | 232 (37.8) | 207 (52.1) | 315 (63.0) |
| Male | 757 (50.1) | 382 (62.2) | 190 (47.9) | 185 (37.0) |
| Townsend deprivation index, mean (SD) | -1.5 (2.9) | -1.5 (2.9) | -1.7 (2.7) | -1.5 (3.0) |
| Body mass index, kg/m^2^, mean (SD) | 27.0 (4.6) | 27.6 (4.4) | 26.9 (4.9) | 26.4 (4.7) |
| Education, n (%) |  |  |  |  |
| College | 577 (38.2) | 195 (31.8) | 155 (39.0) | 227 (45.4) |
| Below college | 934 (61.8) | 419 (68.2) | 242 (61.0) | 273 (54.6) |
| Ethnicity, n (%) |  |  |  |  |
| White | 49 (3.2) | 19 (3.1) | 14 (3.5) | 16 (3.2) |
| Others | 1462 (96.8) | 595 (96.9) | 383 (96.5) | 484 (96.8) |
| Smoking status, n (%) |  |  |  |  |
| Never | 754 (49.9) | 309 (50.3) | 190 (47.9) | 255 (51.0) |
| Previous | 675 (44.7) | 260 (42.3) | 185 (46.6) | 230 (46.0) |
| Current | 82 (5.4) | 45 (7.3) | 22 (5.5) | 15 (3.0) |
| Physical activity, n (%) |  |  |  |  |
| Low | 504 (33.4) | 249 (40.6) | 124 (31.2) | 131 (26.2) |
| Moderate | 478 (31.6) | 170 (27.7) | 132 (33.2) | 176 (35.2) |
| High | 529 (35.0) | 195 (31.8) | 141 (35.5) | 193 (38.6) |
| Total energy, KJ/d, mean (SD) | 8665.6 (2043.3) | 9049.2 (2104.3) | 8573.6 (2013.6) | 8267.4 (1905.2) |
| IBD-specific characteristic |  |  |  |  |
| INFLA score, mean (SD) | 1.2 (6.1) | 1.6 (5.9) | 1.2 (6.0) | 0.7 (6.4) |
| Duration of IBD, years, mean (SD) | 18.2 (13.5) | 18.7 (13.4) | 17.4 (13.3) | 18.4 (13.6) |
| Baseline IBD-related surgery history, n (%) | 170 (11.3) | 83 (13.5) | 47 (11.8) | 40 (8.0) |
| IBD related medication, n (%) | 612 (40.5) | 261 (42.5) | 150 (37.8) | 201 (40.2) |
| With obstruction or stricture, n (%) | 863 (57.1) | 352 (57.3) | 222 (55.9) | 289 (57.8) |
| With fistula or perforation, n (%) | 823 (54.5) | 326 (53.1) | 217 (54.7) | 280 (56.0) |

MIND, Mediterranean-Dietary Approaches to Stop Hypertension Intervention for Neurodegenerative Delay; SD, standard deviation.; IBD, inflammatory bowel disease.

### Table S7. Baseline characteristics of participants with inflammatory bowel disease according to presence or absence of dietary recall records.

| **Characteristic** |  | **Dietary data status** | |
| --- | --- | --- | --- |
|  | **Overall (n=9266)** | **Without dietary data (n=5612)** | **With dietary data (n=3654)** |
| Age, mean (SD) | 57.3 (8.0) | 57.7 (8.0) | 56.6 (8.0) |
| Sex, n (%) |  |  |  |
| Female | 4793 (51.7) | 2955 (52.7) | 1838 (50.3) |
| Male | 4473 (48.3) | 2657 (47.3) | 1816 (49.7) |
| Townsend deprivation index, mean (SD) | -1.1 (3.2) | -1.0 (3.3) | -1.4(3.0) |
| Body mass index, mean (SD) | 27.5 (4.8) | 27.8 (4.9) | 27.1 (4.6) |
| Education, n (%) |  |  |  |
| College | 2481 (27.3) | 1162 (21.3) | 1319 (36.4) |
| Below college | 6594 (72.7) | 4289 (78.7) | 2305 (63.6) |
| Ethnicity，n (%) |  |  |  |
| White | 8794 (95.1) | 5293 (94.6) | 3501 (95.9) |
| Others | 455 (4.9) | 305 (5.4) | 150 (4.1) |
| Smoking status, n (%) |  |  |  |
| Never | 4206 (45.5) | 2450 (43.8) | 1756 (48.1) |
| Previous | 3940 (42.6) | 2341 (41.8) | 1599 (43.8) |
| Current | 1103 (11.9) | 807 (14.4) | 296 (8.1) |
| IBD subtype, n (%) |  |  |  |
| Crohn’s disease | 3196 (34.5) | 1983 (35.3) | 1213 (33.2) |
| Ulcerative colitis | 6070 (65.5) | 3629 (64.7) | 2441 (66.8) |

MIND, Mediterranean-Dietary Approaches to Stop Hypertension Intervention for Neurodegenerative Delay; SD, standard deviation.; IBD, inflammatory bowel disease.

### Table S8. Secondary analysis for associations between individual components of MIND diet and risk of incident IBD-related surgery.

| **Food component** | **Inflammatory bowel disease** | | **Crohn’s disease** | | **Ulcerative colitis** | |
| --- | --- | --- | --- | --- | --- | --- |
|  | **HR (95% CI) ^a^** | ***P-*value** | **HR (95% CI)** | ***P-*value** | **HR (95% CI)** | ***P-*value** |
| Green leafy vegetables | 0.71 (0.47, 1.08) | 0.109 | 0.48 (0.23, 1.02) | 0.057 | 0.93 (0.56, 1.55) | 0.779 |
| Other vegetables | 0.62 (0.40, 0.96) | **0.031** | 0.48 (0.25, 0.92) | **0.027** | 0.75 (0.41, 1.40) | 0.370 |
| Berries | 0.73 (0.48, 1.12) | 0.151 | 0.69 (0.35, 1.36) | 0.284 | 0.81 (0.47, 1.40) | 0.448 |
| Nuts | 0.83 (0.48, 1.45) | 0.520 | 0.51 (0.19, 1.38) | 0.184 | 1.15 (0.58, 2.27) | 0.681 |
| Olive oil | 0.79 (0.58, 1.08) | 0.134 | 0.89 (0.55, 1.44) | 0.632 | 0.73 (0.48, 1.10) | 0.129 |
| Butter/margarine | 0.63 (0.41, 0.96) | **0.031** | 0.43 (0.22, 0.86) | **0.017** | 0.85 (0.49, 1.47) | 0.566 |
| Cheese | 1.00 (0.67, 1.51) | 0.981 | 0.80 (0.43, 1.51) | 0.500 | 1.11 (0.65, 1.89) | 0.708 |
| Whole grains | 0.78 (0.47, 1.28) | 0.327 | 0.44 (0.19, 1.04) | 0.061 | 1.09 (0.58, 2.03) | 0.791 |
| Fish (not fried) | 0.81 (0.58, 1.13) | 0.216 | 0.92 (0.56, 1.53) | 0.759 | 0.76 (0.49, 1.17) | 0.212 |
| Beans | 1.22 (0.82, 1.82) | 0.330 | 1.15 (0.59, 2.23) | 0.683 | 1.29 (0.78, 2.14) | 0.324 |
| Poultry (not fried) | 0.76 (0.54, 1.08) | 0.128 | 0.77 (0.46, 1.32) | 0.344 | 0.76 (0.48, 1.20) | 0.238 |
| Red meat and products | 1.01 (0.73, 1.39) | 0.948 | 1.05 (0.64, 1.73) | 0.854 | 1.02 (0.67, 1.56) | 0.916 |
| Fast/fried foods | 0.49 (0.34, 0.70) | **<0.001** | 0.48 (0.27, 0.83) | **0.009** | 0.50 (0.32, 0.80) | **0.004** |
| Pastries and sweets | 0.59 (0.31, 1.11) | 0.100 | 0.52 (0.18, 1.54) | 0.239 | 0.66 (0.30, 1.45) | 0.304 |
| Wine | 1.05 (0.67, 1.66) | 0.834 | 0.88 (0.42, 1.86) | 0.745 | 1.19 (0.66, 2.12) | 0.566 |

a. Model adjusted for age, sex, ethnicity, education, BMI, TDI, smoking status, physical activity, and total energy.

### Table S9. Secondary analysis for associations between MIND diet score tertiles and risk of incident IBD-related surgery stratified by surgical site.

| **MIND diet score** | **Inflammatory bowel disease** | | | **Crohn’s disease** | | | **Ulcerative colitis** | | |
| --- | --- | --- | --- | --- | --- | --- | --- | --- | --- |
|  | **Cases/person-years** | **HR (95% CI) ^a^** | ***P-*value** | **Cases/person-years** | **HR (95% CI)** | ***P-*value** | **Cases/person-years** | **HR (95% CI)** | ***P-*value** |
| **Small bowel resection surgery** | | | | | | | | | |
| Per 3-point increment |  | 0.60 (0.42, 0.85) | **0.004** |  | 0.55 (0.32, 0.94) | **0.029** |  | 0.66 (0.42, 1.04) | 0.072 |
| Tertile 1 | 47/9401 | 1 (Reference) |  | 20/2597 | 1 (Reference) |  | 26/6105 | 1 (Reference) |  |
| Tertile 2 | 19/6006 | 0.69 (0.40, 1.18) | 0.178 | 11/2750 | 0.55 (0.26, 1.18) | 0.124 | 9/3955 | 0.64 (0.30, 1.38) | 0.254 |
| Tertile 3 | 21/7190 | 0.69 (0.40, 1.18) | 0.177 | 7/2140 | 0.46 (0.19, 1.12) | 0.087 | 14/5049 | 0.86 (0.43, 1.71) | 0.666 |
| *P*-trend |  |  | 0.138 |  |  | 0.080 |  |  | 0.574 |
| **Colorectal resection surgery** | | | | | | | | | |
| Per 3-point increment |  | 0.65 (0.46, 0.93) | **0.018** |  | 0.45 (0.25, 0.81) | **0.008** |  | 0.84 (0.54, 1.30) | 0.427 |
| Tertile 1 | 44/9398 | 1 (Reference) |  | 16/2560 | 1 (Reference) |  | 25/6133 | 1 (Reference) |  |
| Tertile 2 | 19/5960 | 0.69 (0.40, 1.19) | 0.185 | 10/2681 | 0.53 (0.23, 1.19) | 0.123 | 12/3983 | 0.87 (0.43, 1.75) | 0.692 |
| Tertile 3 | 21/7264 | 0.63 (0.37, 1.09) | 0.098 | 7/2177 | 0.38 (0.15, 0.95) | **0.039** | 14/5087 | 0.85 (0.43, 1.69) | 0.650 |
| *P*-trend |  |  | 0.080 |  |  | **0.042** |  |  | 0.633 |
| **Proctological surgery** | | | | | | | | | |
| Per 3-point increment |  | 0.47 (0.29, 0.77) | **0.003** |  | 0.47 (0.22, 1.00) | 0.051 |  | 0.47 (0.24, 0.92) | **0.028** |
| Tertile 1 | 30/9130 | 1 (Reference) |  | 10/2499 | 1 (Reference) |  | 16/5930 | 1 (Reference) |  |
| Tertile 2 | 7/5847 | 0.36 (0.16, 0.83) | **0.017** | 5/2639 | 0.40 (0.13, 1.19) | 0.099 | 6/3909 | 0.59 (0.23, 1.54) | 0.283 |
| Tertile 3 | 10/7118 | 0.44 (0.21, 0.92) | **0.029** | 5/2147 | 0.50 (0.16, 1.52) | 0.222 | 5/4971 | 0.41 (0.14, 1.16) | 0.093 |
| *P*-trend |  |  | **0.013** |  |  | 0.244 |  |  | 0.078 |

a． Model adjusted for age, sex, ethnicity, education, BMI, TDI, smoking status, physical activity, and total energy.

### Table S10. Secondary analysis for associations between MIND diet score tertiles and risk of incident IBD-related surgery stratified by hospital admission source.

| **MIND diet score** |  | **Inflammatory bowel disease** | |  | **Crohn’s disease** | |  | **Ulcerative colitis** | |
| --- | --- | --- | --- | --- | --- | --- | --- | --- | --- |
|  | **Cases/person-years** | **HR (95% CI) ^a^** | ***P-*value** | **Cases/person-years** | **HR (95% CI)** | ***P-*value** | **Cases/person-years** | **HR (95% CI)** | ***P-*value** |
| Elective surgery | | | | | | | | | |
| Per 3-point increment |  | 0.64 (0.48, 0.86) | **0.003** |  | 0.60 (0.37, 0.97) | **0.037** |  | 0.69 (0.48, 1.01) | 0.054 |
| Tertile 1 | 65/9551 | 1 (Reference) |  | 22/2650 | 1 (Reference) |  | 39/6179 | 1 (Reference) |  |
| Tertile 2 | 25/6100 | 0.61 (0.38, 0.97) | **0.038** | 14/2809 | 0.56 (0.29, 1.12) | 0.101 | 15/4013 | 0.64 (0.35, 1.17) | 0.146 |
| Tertile 3 | 30/7318 | 0.61 (0.39, 0.96) | **0.031** | 11/2200 | 0.52 (0.25, 1.11) | 0.091 | 19/5118 | 0.66 (0.37, 1.17) | 0.153 |
| *P*-trend |  |  | **0.020** |  |  | 0.099 |  |  | 0.124 |
| Emergency surgery | | | | | | | | | |
| Per 3-point increment |  | 0.56 (0.35, 0.91) | **0.020** |  | 0.37 (0.18, 0.76) | **0.007** |  | 0.88 (0.46, 1.67) | 0.697 |
| Tertile 1 | 23/9133 | 1 (Reference) |  | 13/2493 | 1 (Reference) |  | 10/5954 | 1 (Reference) |  |
| Tertile 2 | 12/5868 | 0.85 (0.42, 1.73) | 0.658 | 6/2631 | 0.43 (0.16, 1.16) | 0.097 | 6/3923 | 1.07 (0.38, 2.99) | 0.896 |
| Tertile 3 | 11/7140 | 0.71 (0.34, 1.49) | 0.359 | 3/2135 | 0.26 (0.07, 0.94) | **0.041** | 8/5005 | 1.32 (0.50, 3.46) | 0.578 |
| *P*-trend |  |  | 0.354 |  |  | **0.032** |  |  | 0.586 |

a． Model adjusted for age, sex, ethnicity, education, BMI, TDI, smoking status, physical activity, and total energy.

### Table S11. Subgroup analysis for associations between tertiles of MIND diet score and the risk of incident IBD-related surgery stratified by age, sex, and body mass index.

| **MIND score** | **Inflammatory bowel disease** | | **Crohn’s disease** | | **Ulcerative colitis** | |
| --- | --- | --- | --- | --- | --- | --- |
|  | **HR (95% CI) ^a^** | ***P*-value** | **HR (95% CI)** | ***P*-value** | **HR (95% CI)** | ***P*-value** |
| **Age** | | | | | | |
| **Age>=60** |  |  |  |  |  |  |
| Per 3-point increment | 0.71 (0.49, 1.02) | 0.066 | 0.55 (0.28, 1.07) | 0.076 | 0.80 (0.51, 1.27) | 0.349 |
| Tertile 1 | 1 (Reference) |  | 1 (Reference) |  | 1 (Reference) |  |
| Tertile 2 | 0.50 (0.26, 0.95) | **0.034** | 0.33 (0.12, 0.94) | **0.038** | 0.51 (0.23, 1.14) | 0.103 |
| Tertile 3 | 0.79 (0.46, 1.36) | 0.400 | 0.55 (0.21, 1.43) | 0.219 | 0.86 (0.44, 1.66) | 0.650 |
| *P*-trend |  | 0.274 |  | 0.242 |  | 0.528 |
| **Age<60** |  |  |  |  |  |  |
| Per 3-point increment | 0.58 (0.41, 0.81) | **0.002** | 0.52 (0.31, 0.87) | **0.012** | 0.71 (0.45, 1.11) | 0.134 |
| Tertile 1 | 1 (Reference) |  | 1 (Reference) |  | 1 (Reference) |  |
| Tertile 2 | 0.82 (0.50, 1.35) | 0.443 | 0.69 (0.35, 1.35) | 0.280 | 1.03 (0.51, 2.08) | 0.928 |
| Tertile 3 | 0.54 (0.31, 0.94) | **0.030** | 0.40 (0.17, 0.97) | **0.043** | 0.76 (0.37, 1.60) | 0.473 |
| *P*-trend |  | **0.031** |  | **0.040** |  | 0.502 |
| *P-*interaction ^b^ |  | 0.163 |  | 0.320 |  | 0.366 |
| **Sex** | | | | | | |
| **Female** |  |  |  |  |  |  |
| Per 3-point increment | 0.73 (0.51, 1.04) | 0.084 | 0.47 (0.27, 0.80) | **0.005** | 1.17 (0.71, 1.92) | 0.545 |
| Tertile 1 | 1 (Reference) |  | 1 (Reference) |  | 1 (Reference) |  |
| Tertile 2 | 0.93 (0.54, 1.59) | 0.786 | 0.75 (0.37, 1.52) | 0.427 | 1.02 (0.44, 2.38) | 0.966 |
| Tertile 3 | 0.63 (0.36, 1.09) | 0.098 | 0.31 (0.12, 0.77) | **0.012** | 1.05 (0.48, 2.28) | 0.900 |
| *P*-trend |  | 0.100 |  | **0.010** |  | 0.899 |
| **Male** |  |  |  |  |  |  |
| Per 3-point increment | 0.54 (0.38, 0.78) | **0.001** | 0.57 (0.30, 1.06) | 0.077 | 0.55 (0.35, 0.85) | **0.007** |
| Tertile 1 | 1 (Reference) |  | 1 (Reference) |  | 1 (Reference) |  |
| Tertile 2 | 0.51 (0.28, 0.92) | **0.025** | 0.24 (0.08, 0.72) | **0.011** | 0.65 (0.33, 1.29) | 0.223 |
| Tertile 3 | 0.68 (0.39, 1.16) | 0.155 | 0.74 (0.30, 1.86) | 0.525 | 0.65 (0.33, 1.29) | 0.216 |
| *P*-trend |  | 0.066 |  | 0.402 |  | 0.155 |
| *P-*interaction |  | 0.232 |  | **0.035** |  | 0.602 |
| **Body mass index (BMI)** | | | | | | |
| **BMI>= 30** |  |  |  |  |  |  |
| Per 3-point increment | 0.67 (0.39, 1.16) | 0.154 | 0.27 (0.07, 1.09) | 0.065 | 0.79 (0.42, 1.48) | 0.467 |
| Tertile 1 | 1 (Reference) |  | 1 (Reference) |  | 1 (Reference) |  |
| Tertile 2 | 0.80 (0.35, 1.86) | 0.609 | 0.42 (0.08, 2.15) | 0.300 | 0.94 (0.33, 2.72) | 0.915 |
| Tertile 3 | 0.66 (0.28, 1.56) | 0.341 | 0.17 (0.02, 1.79) | 0.140 | 0.91 (0.34, 2.43) | 0.849 |
| *P*-trend |  | 0.329 |  | 0.136 |  | 0.845 |
| **BMI< 30** |  |  |  |  |  |  |
| Per 3-point increment | 0.64 (0.48, 0.85) | **0.002** | 0.56 (0.36, 0.86) | **0.008** | 0.75 (0.52, 1.08) | 0.125 |
| Tertile 1 | 1 (Reference) |  | 1 (Reference) |  | 1 (Reference) |  |
| Tertile 2 | 0.69 (0.44, 1.06) | 0.092 | 0.51 (0.28, 0.96) | **0.036** | 0.72 (0.40, 1.31) | 0.285 |
| Tertile 3 | 0.66 (0.43, 1.01) | 0.056 | 0.51 (0.26, 1.01) | 0.052 | 0.75 (0.42, 1.32) | 0.319 |
| *P*-trend |  | **0.041** |  | 0.054 |  | 0.285 |
| *P-*interaction |  | 0.798 |  | 0.785 |  | 0.638 |

MIND, Mediterranean-Dietary Approaches to Stop Hypertension Intervention for Neurodegenerative Delay; HR, hazard ratios; CI, confidence interval.

a. Adjusted for all covariates in Model 2 except for the one defining subgroup.
b. Multiplicative interactions were tested by adding multiplicative interaction terms into the model.

### Table S12. Subgroup analysis for associations between tertiles of MIND diet score and the risk of incident IBD-related surgery stratified by INFLA score, duration of IBD before baseline, and IBD-related surgery before baseline.

| **MIND score** | **Inflammatory bowel disease** | | **Crohn’s disease** | | **Ulcerative colitis** | |
| --- | --- | --- | --- | --- | --- | --- |
|  | **HR (95% CI) ^a^** | ***P*-value** | **HR (95% CI)** | ***P*-value** | **HR (95% CI)** | ***P*-value** |
| **Inflammation severity** | | | | | | |
| **High (INFLA>0)** |  |  |  |  |  |  |
| Per 3-point increment | 0.62 (0.46, 0.83) | **0.002** | 0.59 (0.38, 0.94) | **0.027** | 0.66 (0.45, 0.97) | **0.037** |
| Tertile 1 | 1 (Reference) |  | 1 (Reference) |  | 1 (Reference) |  |
| Tertile 2 | 0.68 (0.43, 1.07) | 0.094 | 0.54 (0.29, 1.02) | 0.058 | 0.69 (0.37, 1.28) | 0.240 |
| Tertile 3 | 0.61 (0.38, 0.97) | **0.039** | 0.54 (0.26, 1.12) | 0.099 | 0.63 (0.34, 1.17) | 0.145 |
| *P*-trend |  | **0.025** |  | 0.084 |  | 0.118 |
| **Low (INFLA<=0)** |  |  |  |  |  |  |
| Per 3-point increment | 0.71 (0.44, 1.14) | 0.154 | 0.26 (0.09, 0.74) | **0.011** | 0.97 (0.55, 1.71) | 0.914 |
| Tertile 1 | 1 (Reference) |  | 1 (Reference) |  | 1 (Reference) |  |
| Tertile 2 | 0.73 (0.34, 1.59) | 0.431 | 0.33 (0.09, 1.26) | 0.104 | 0.98 (0.37, 2.56) | 0.960 |
| Tertile 3 | 0.80 (0.39, 1.61) | 0.523 | 0.21 (0.05, 0.94) | **0.041** | 1.19 (0.51, 2.82) | 0.685 |
| *P*-trend |  | 0.512 |  | 0.056 |  | 0.683 |
| *P-*interaction ^b^ |  | 0.230 |  | 0.956 |  | 0.089 |
| **Duration of IBD** | | | | | | |
| **Duration < =15 years** |  |  |  |  |  |  |
| Per 3-point increment | 0.48 (0.33, 0.71) | **<0.001** | 0.33 (0.16, 0.69) | **0.003** | 0.56 (0.35, 0.89) | **0.015** |
| Tertile 1 | 1 (Reference) |  | 1 (Reference) |  | 1 (Reference) |  |
| Tertile 2 | 0.90 (0.54, 1.52) | 0.702 | 0.58 (0.24, 1.37) | 0.212 | 1.03 (0.53, 2.00) | 0.923 |
| Tertile 3 | 0.46 (0.24, 0.88) | **0.018** | 0.29 (0.09, 0.90) | **0.032** | 0.56 (0.25, 1.23) | 0.150 |
| *P*-trend |  | **0.023** |  | **0.028** |  | 0.184 |
| **Duration > 15 years** |  |  |  |  |  |  |
| Per 3-point increment | 0.76 (0.55, 1.06) | 0.107 | 0.61 (0.37, 1.01) | 0.053 | 0.98 (0.63, 1.53) | 0.934 |
| Tertile 1 | 1 (Reference) |  | 1 (Reference) |  | 1 (Reference) |  |
| Tertile 2 | 0.49 (0.27, 0.89) | **0.018** | 0.49 (0.23, 1.06) | 0.069 | 0.44 (0.18, 1.09) | 0.078 |
| Tertile 3 | 0.77 (0.47, 1.26) | 0.298 | 0.55 (0.25, 1.22) | 0.144 | 1.02 (0.53, 1.96) | 0.960 |
| *P*-trend |  | 0.206 |  | 0.161 |  | 0.928 |
| *P-*interaction |  | **0.047** |  | 0.541 |  | 0.061 |
| **IBD-related surgery at baseline** | | | | | | |
| **Free of surgery** |  |  |  |  |  |  |
| Per 3-point increment | 0.64 (0.47, 0.86) | **0.003** | 0.55 (0.33, 0.91) | **0.019** | 0.71 (0.49, 1.03) | 0.072 |
| Tertile 1 | 1 (Reference) |  | 1 (Reference) |  | 1 (Reference) |  |
| Tertile 2 | 0.65 (0.41, 1.03) | 0.064 | 0.72 (0.36, 1.43) | 0.344 | 0.53 (0.28, 1.00) | 0.052 |
| Tertile 3 | 0.63 (0.40, 0.97) | **0.038** | 0.40 (0.18, 0.91) | **0.029** | 0.76 (0.44, 1.30) | 0.309 |
| *P*-trend |  | **0.028** |  | **0.029** |  | 0.242 |
| **With surgery history** |  |  |  |  |  |  |
| Per 3-point increment | 0.67 (0.42, 1.05) | 0.083 | 0.53 (0.28, 1.01) | 0.055 | 0.90 (0.47, 1.75) | 0.762 |
| Tertile 1 | 1 (Reference) |  | 1 (Reference) |  | 1 (Reference) |  |
| Tertile 2 | 0.88 (0.43, 1.81) | 0.733 | 0.25 (0.08, 0.76) | **0.015** | 1.97 (0.69, 5.60) | 0.204 |
| Tertile 3 | 0.78 (0.35, 1.73) | 0.548 | 0.65 (0.22, 1.91) | 0.434 | 0.89 (0.25, 3.15) | 0.861 |
| *P*-trend |  | 0.533 |  | 0.276 |  | 0.968 |
| *P-*interaction |  | 0.843 |  | 0.091 |  | 0.114 |

MIND, Mediterranean-Dietary Approaches to Stop Hypertension Intervention for Neurodegenerative Delay; HR, hazard ratios; CI, confidence interval.

a. Adjusted for all covariates in Model 2 except for the one defining subgroup.
b. Multiplicative interactions were tested by adding multiplicative interaction terms into the model.

### Table S13. Subgroup analysis for associations between tertiles of MIND diet score and the risk of incident IBD-related surgery stratified by IBD-related medication, obstruction or stricture status, and fistula or perforation status.

| **MIND score** | **Inflammatory bowel disease** | | **Crohn’s disease** | | **Ulcerative colitis** | |
| --- | --- | --- | --- | --- | --- | --- |
|  | **HR (95% CI) ^a^** | ***P*-value** | **HR (95% CI)** | ***P*-value** | **HR (95% CI)** | ***P*-value** |
| **IBD-related medication** | | | | | | |
| **Non-users of IBD-related medications** |  |  |  |  |  |  |
| Per 3-point increment | 0.59 (0.42, 0.83) | **0.003** | 0.53 (0.31, 0.89) | **0.017** | 0.67 (0.43, 1.06) | 0.085 |
| Tertile 1 | 1 (Reference) |  | 1 (Reference) |  | 1 (Reference) |  |
| Tertile 2 | 0.76 (0.45, 1.25) | 0.278 | 0.56 (0.27, 1.17) | 0.125 | 0.76 (0.37, 1.52) | 0.433 |
| Tertile 3 | 0.65 (0.39, 1.10) | 0.109 | 0.47 (0.21, 1.07) | 0.071 | 0.75 (0.38, 1.50) | 0.420 |
| *P*-trend |  | 0.097 |  | 0.080 |  | 0.393 |
| **Users of IBD-related medications** |  |  |  |  |  |  |
| Per 3-point increment | 0.68 (0.47, 0.99) | **0.043** | 0.48 (0.25, 0.92) | **0.026** | 0.86 (0.54, 1.35) | 0.507 |
| Tertile 1 | 1 (Reference) |  | 1 (Reference) |  | 1 (Reference) |  |
| Tertile 2 | 0.62 (0.34, 1.13) | 0.120 | 0.44 (0.18, 1.11) | 0.081 | 0.77 (0.35, 1.68) | 0.511 |
| Tertile 3 | 0.65 (0.36, 1.15) | 0.136 | 0.37 (0.13, 1.07) | 0.066 | 0.87 (0.43, 1.78) | 0.712 |
| *P*-trend |  | 0.097 |  | 0.060 |  | 0.667 |
| *P-*interaction ^b^ |  | 0.856 |  | 0.821 |  | 0.978 |
| **Obstruction or stricture** | | | | | | |
| **Without obstruction and stricture** |  |  |  |  |  |  |
| Per 3-point increment | 0.99 (0.63, 1.55) | 0.964 | 0.67 (0.33, 1.34) | 0.255 | 1.43 (0.79, 2.59) | 0.233 |
| Tertile 1 | 1 (Reference) |  | 1 (Reference) |  | 1 (Reference) |  |
| Tertile 2 | 1.64 (0.83, 3.26) | 0.158 | 1.03 (0.38, 2.80) | 0.949 | 2.24 (0.84, 5.93) | 0.105 |
| Tertile 3 | 1.32 (0.65, 2.67) | 0.437 | 0.59 (0.18, 1.86) | 0.366 | 2.52 (0.97, 6.53) | 0.057 |
| *P*-trend |  | 0.393 |  | 0.351 |  | 0.054 |
| **With obstruction or stricture** |  |  |  |  |  |  |
| Per 3-point increment | 0.52 (0.39, 0.71) | **<0.001** | 0.47 (0.28, 0.80) | **0.005** | 0.59 (0.40, 0.86) | **0.006** |
| Tertile 1 | 1 (Reference) |  | 1 (Reference) |  | 1 (Reference) |  |
| Tertile 2 | 0.47 (0.29, 0.77) | **0.003** | 0.38 (0.19, 0.77) | **0.007** | 0.50 (0.26, 0.97) | **0.040** |
| Tertile 3 | 0.47 (0.29, 0.76) | **0.002** | 0.45 (0.21, 1.00) | **0.049** | 0.51 (0.28, 0.94) | **0.030** |
| *P*-trend |  | **0.001** |  | **0.039** |  | **0.018** |
| *P-*interaction |  | **0.005** |  | 0.257 |  | **0.020** |
| **Fistula or perforation** | | | | | | |
| **Without fistula or perforation** |  |  |  |  |  |  |
| Per 3-point increment | 0.74 (0.52, 1.05) | 0.090 | 0.71 (0.43, 1.18) | 0.185 | 0.81 (0.50, 1.33) | 0.408 |
| Tertile 1 | 1 (Reference) |  | 1 (Reference) |  | 1 (Reference) |  |
| Tertile 2 | 1.19 (0.71, 1.99) | 0.510 | 0.85 (0.41, 1.78) | 0.675 | 1.62 (0.78, 3.38) | 0.196 |
| Tertile 3 | 0.83 (0.48, 1.43) | 0.503 | 0.64 (0.29, 1.43) | 0.279 | 1.10 (0.49, 2.43) | 0.821 |
| *P*-trend |  | 0.574 |  | 0.277 |  | 0.722 |
| **With fistula or perforation** |  |  |  |  |  |  |
| Per 3-point increment | 0.54 (0.38, 0.77) | **0.001** | 0.32 (0.16, 0.64) | **0.001** | 0.69 (0.45, 1.06) | 0.089 |
| Tertile 1 | 1 (Reference) |  | 1 (Reference) |  | 1 (Reference) |  |
| Tertile 2 | 0.35 (0.19, 0.67) | **0.001** | 0.27 (0.10, 0.69) | **0.007** | 0.34 (0.15, 0.77) | **0.010** |
| Tertile 3 | 0.50 (0.29, 0.87) | **0.014** | 0.24 (0.07, 0.86) | **0.028** | 0.62 (0.33, 1.18) | 0.144 |
| *P*-trend |  | **0.004** |  | **0.010** |  | 0.088 |
| *P-*interaction |  | **0.005** |  | **0.022** |  | **0.024** |

MIND, Mediterranean-Dietary Approaches to Stop Hypertension Intervention for Neurodegenerative Delay; HR, hazard ratios; CI, confidence interval.

a. Adjusted for all covariates in Model 2 except for the one defining subgroup.
b. Multiplicative interactions were tested by adding multiplicative interaction terms into the model.

### Table S14. Additive Interaction of MIND diet score and baseline characteristics on the risk of incident IBD-related surgery

|  | **RERI (95%CI) ^a^** | ***P-*value** | **AP (95%CI)** | ***P-*value** | **SI (95%CI)** | ***P-*value** |
| --- | --- | --- | --- | --- | --- | --- |
| Inflammatory bowel disease | | | | | | |
| Age | -0.45 (-1.69, 0.78) | 0.764 | -0.22 (-0.83, 0.38) | 0.234 | 0.69 (0.3, 1.63) | 0.800 |
| Sex | 0.07 (-1, 1.14) | 0.448 | 0.03 (-0.49, 0.56) | 0.448 | 1.07 (0.36, 3.19) | 0.450 |
| BMI | -0.41 (-1.77, 0.95) | 0.724 | -0.17 (-0.74, 0.39) | 0.270 | 0.77 (0.36, 1.62) | 0.756 |
| INFLA score | 0.8 (-0.14, 1.73) | **0.047** | 0.34 (-0.07, 0.75) | 0.054 | 2.44 (0.32, 18.5) | 0.193 |
| Duration of IBD | -0.69 (-2.28, 0.91) | 0.801 | -0.25 (-0.82, 0.31) | 0.188 | 0.71 (0.38, 1.35) | 0.849 |
| With surgery history | 0.16 (-1.9, 2.22) | 0.439 | 0.05 (-0.59, 0.69) | 0.438 | 1.08 (0.4, 2.93) | 0.440 |
| With IBD-related medication use | 0.25 (-0.73, 1.23) | 0.309 | 0.12 (-0.36, 0.61) | 0.308 | 1.33 (0.36, 4.96) | 0.334 |
| With obstruction or stricture ^b^ | - | - | - | - | - | - |
| With fistula or perforation | 1.08 (0.21, 1.95) | **0.008** | 0.47 (0.12, 0.82) | **0.004** | 5.79 (0.07, 452.62) | 0.215 |
| Crohn’s disease | | | | | | |
| Age | -0.47 (-2.69, 1.75) | 0.662 | -0.2 (-1.15, 0.75) | 0.340 | 0.74 (0.21, 2.62) | 0.679 |
| Sex | -0.88 (-3.48, 1.71) | 0.748 | -0.34 (-1.33, 0.64) | 0.248 | 0.64 (0.22, 1.86) | 0.792 |
| BMI | -0.63 (-3.64, 2.37) | 0.660 | -0.18 (-1, 0.64) | 0.335 | 0.8 (0.32, 2.01) | 0.682 |
| INFLA score | 0.53 (-2.3, 3.37) | 0.356 | 0.12 (-0.55, 0.8) | 0.359 | 1.19 (0.41, 3.45) | 0.373 |
| Duration of IBD | -0.49 (-3.52, 2.53) | 0.625 | -0.13 (-0.91, 0.65) | 0.372 | 0.85 (0.34, 2.1) | 0.639 |
| With surgery history | 1.29 (-2.43, 5.02) | 0.248 | 0.25 (-0.4, 0.9) | 0.227 | 1.45 (0.45, 4.63) | 0.266 |
| With IBD-related medication use | -0.3 (-2.37, 1.76) | 0.613 | -0.13 (-1.02, 0.76) | 0.384 | 0.81 (0.22, 2.9) | 0.629 |
| With obstruction or stricture | - | - | - | - | - | - |
| With fistula or perforation | 3.4 (0.52, 6.29) | **0.010** | 0.72 (0.39, 1.06) | **<0.001** | 11.95 (0.04, 3448.73) | 0.195 |
| Ulcerative colitis | | | | | | |
| Age | -0.18 (-1.67, 1.3) | 0.596 | -0.09 (-0.8, 0.62) | 0.404 | 0.85 (0.26, 2.77) | 0.603 |
| Sex | 0.73 (-0.4, 1.86) | 0.102 | 0.37 (-0.2, 0.94) | 0.100 | 3.99 (0.02, 712.33) | 0.300 |
| BMI | -0.04 (-1.37, 1.28) | 0.525 | -0.02 (-0.73, 0.69) | 0.475 | 0.95 (0.22, 4.11) | 0.526 |
| INFLA score | - | - | - | - | - | - |
| Duration of IBD | -0.94 (-2.95, 1.06) | 0.822 | -0.42 (-1.26, 0.43) | 0.166 | 0.57 (0.23, 1.43) | 0.885 |
| With surgery history | -0.3 (-2.86, 2.26) | 0.591 | -0.13 (-1.32, 1.05) | 0.412 | 0.8 (0.14, 4.78) | 0.595 |
| With IBD-related medication use | 0.14 (-1.2, 1.48) | 0.420 | 0.07 (-0.59, 0.72) | 0.419 | 1.15 (0.26, 5.05) | 0.425 |
| With obstruction or stricture | - | - | - | - | - | - |
| With fistula or perforation | 0.68 (-0.38, 1.75) | 0.104 | 0.34 (-0.2, 0.89) | 0.108 | 3.24 (0.05, 221.53) | 0.293 |

MIND, Mediterranean-Dietary Approaches to Stop Hypertension Intervention for Neurodegenerative Delay; RERI, the relative excess risk due to interaction; AP, attributable proportion due to interaction; SI, synergy index; CI, confidence interval. Positive RERI and AP, with SI > 1, indicate additive synergism..

a. Model additionally adjusted for age, sex, ethnicity, education, BMI, TDI, smoking status, physical activity, and total energy.
b. The absence of detectable additive interaction likely reflects insufficient sample size and unstable estimates.

### Table S15. Sensitivity analysis for associations between quartiles of MIND diet score and the risk of incident IBD-related surgery.

| **MIND diet score** | **Inflammatory bowel disease** | | **Crohn’s disease** | | **Ulcerative colitis** | |
| --- | --- | --- | --- | --- | --- | --- |
|  | **HR (95% CI) ^a^** | ***P-*value** | **HR (95% CI)** | ***P-*value** | **HR (95% CI)** | ***P-*value** |
| Per 3-point increment | 0.63 (0.49, 0.81) | **<0.001** | 0.52 (0.35, 0.78) | **0.001** | 0.75 (0.54, 1.03) | 0.076 |
| Quartile 1 | 1 (Reference) |  | 1 (Reference) |  | 1 (Reference) |  |
| Quartile 2 | 0.50 (0.31, 0.81) | **0.005** | 0.41 (0.19, 0.91) | **0.028** | 0.61 (0.36, 1.04) | 0.069 |
| Quartile 3 | 0.70 (0.48, 1.03) | 0.073 | 0.66 (0.37, 1.18) | 0.165 | 0.83 (0.49, 1.42) | 0.498 |
| Quartile 4 | 0.49 (0.31, 0.78) | **0.002** | 0.33 (0.15, 0.73) | **0.006** | 0.64 (0.33, 1.22) | 0.174 |
| *P*-trend |  | **0.003** |  | **0.011** |  | 0.183 |

MIND, Mediterranean-Dietary Approaches to Stop Hypertension Intervention for Neurodegenerative Delay; HR, hazard ratios; CI, confidence interval.

a. Model adjusted for age, sex, ethnicity, education, BMI, TDI, smoking status, physical activity, and total energy.

### Table S16. Sensitivity analysis for associations between MIND diet score and the risk of incident IBD-related surgery in IBD, CD, and UC according to further adjustment.

| **MIND diet score** | **Inflammatory bowel disease** | | **Crohn’s disease** | | **Ulcerative colitis** | |
| --- | --- | --- | --- | --- | --- | --- |
|  | **HR (95% CI)** | ***P-*value** | **HR (95% CI)** | ***P-*value** | **HR (95% CI)** | ***P-*value** |
| **Excluding new cases in the first years of follow-up** | | | | | | |
| Per 3-point increment | 0.66 (0.51, 0.86) | **0.002** | 0.53 (0.35, 0.81) | **0.003** | 0.80 (0.57, 1.12) | 0.199 |
| Tertile 1 | 1 (Reference) |  | 1 (Reference) |  | 1 (Reference) |  |
| Tertile 2 | 0.66 (0.43, 1.00) | 0.052 | 0.51 (0.28, 0.93) | **0.029** | 0.69 (0.39, 1.22) | 0.206 |
| Tertile 3 | 0.70 (0.47, 1.05) | 0.083 | 0.44 (0.22, 0.86) | **0.017** | 0.90 (0.54, 1.50) | 0.690 |
| *P*-trend |  | 0.058 |  | **0.017** |  | 0.618 |
| **Additionally adjusted for Charlson comorbidity index** | | | | | | |
| Per 3-point increment | 0.63 (0.49, 0.81) | **<0.001** | 0.53 (0.35, 0.79) | **0.002** | 0.75 (0.54, 1.03) | 0.075 |
| Tertile 1 | 1 (Reference) |  | 1 (Reference) |  | 1 (Reference) |  |
| Tertile 2 | 0.68 (0.46, 1.00) | 0.050 | 0.54 (0.31, 0.95) | **0.031** | 0.74 (0.44, 1.24) | 0.251 |
| Tertile 3 | 0.64 (0.44, 0.94) | **0.024** | 0.45 (0.24, 0.86) | **0.016** | 0.79 (0.48, 1.30) | 0.357 |
| *P*-trend |  | **0.016** |  | **0.016** |  | 0.313 |
| **Additionally adjusted for fiber intake** | | | | | | |
| Per 3-point increment | 0.64 (0.49, 0.83) | **0.001** | 0.55 (0.36, 0.84) | **0.005** | 0.75 (0.53, 1.04) | 0.088 |
| Tertile 1 | 1 (Reference) |  | 1 (Reference) |  | 1 (Reference) |  |
| Tertile 2 | 0.70 (0.47, 1.04) | 0.076 | 0.55 (0.31, 0.97) | **0.040** | 0.75 (0.44, 1.26) | 0.272 |
| Tertile 3 | 0.68 (0.45, 1.01) | 0.059 | 0.49 (0.25, 0.97) | **0.040** | 0.81 (0.48, 1.35) | 0.419 |
| *P*-trend |  | **0.042** |  | **0.038** |  | 0.369 |
| **Using a modified wine-excluded MIND diet score** | | | | | | |
| Per 3-point increment | 0.61 (0.47, 0.79) | **<0.001** | 0.50 (0.33, 0.76) | **0.001** | 0.72 (0.52, 1.01) | 0.055 |
| Tertile 1 | 1 (Reference) |  | 1 (Reference) |  | 1 (Reference) |  |
| Tertile 2 | 0.56 (0.39, 0.81) | **0.002** | 0.50 (0.27, 0.91) | **0.023** | 0.64 (0.40, 1.03) | 0.065 |
| Tertile 3 | 0.51 (0.34, 0.77) | **0.001** | 0.40 (0.22, 0.74) | **0.003** | 0.70 (0.42, 1.17) | 0.175 |
| *P*-trend |  | **0.001** |  | **0.002** |  | 0.147 |
| **Using multiple imputation for missing data** | | | | | | |
| Per 3-point increment | 0.64(0.49, 0.82) | **0.001** | 0.52(0.35, 0.79) | **0.003** | 0.75(0.54, 1.03) | 0.082 |
| Tertile 1 | 1 (Reference) |  | 1 (Reference) |  | 1 (Reference) |  |
| Tertile 2 | 0.69(0.47, 1.02) | 0.064 | 0.55(0.31, 0.97) | **0.043** | 0.73(0.44, 1.23) | 0.244 |
| Tertile 3 | 0.65(0.44, 0.96) | **0.030** | 0.45(0.23, 0.86) | **0.019** | 0.79(0.48, 1.30) | 0.359 |
| *P*-trend |  | **0.021** |  | **0.019** |  | 0.313 |
| **Limited to participants with stricter IBD diagnostic criteria** | | | | | | |
| Per 3-point increment | 0.63 (0.48, 0.83) | **0.001** | 0.56 (0.37, 0.83) | **0.004** | 0.74 (0.51, 1.06) | 0.102 |
| Tertile 1 | 1 (Reference) |  | 1 (Reference) |  | 1 (Reference) |  |
| Tertile 2 | 0.71 (0.47, 1.08) | 0.113 | 0.56 (0.32, 0.99) | **0.046** | 0.73 (0.40, 1.32) | 0.298 |
| Tertile 3 | 0.64 (0.42, 0.98) | **0.041** | 0.45 (0.23, 0.87) | **0.019** | 0.81 (0.46, 1.42) | 0.461 |
| *P*-trend |  | **0.030** |  | **0.018** |  | 0.401 |
| **Using competitive risk model** | | | | | | |
| Per 3-point increment | 0.63 (0.48, 0.83) | **<0.001** | 0.52 (0.35, 0.77) | **0.001** | 0.75 (0.52, 1.07) | 0.109 |
| Tertile 1 | 1 (Reference) |  | 1 (Reference) |  | 1 (Reference) |  |
| Tertile 2 | 0.68 (0.46, 0.99) | **0.047** | 0.52 (0.30, 0.90) | **0.020** | 0.73 (0.43, 1.23) | 0.242 |
| Tertile 3 | 0.65 (0.45, 0.94) | **0.024** | 0.44 (0.24, 0.82) | **0.010** | 0.80 (0.50, 1.29) | 0.353 |
| *P*-trend |  | **0.017** |  | **0.014** |  | 0.315 |

MIND, Mediterranean-Dietary Approaches to Stop Hypertension Intervention for Neurodegenerative Delay; HR, hazard ratios; CI, confidence interval.

a. Model adjusted for age, sex, ethnicity, education, BMI, TDI, smoking status, physical activity, and total energy.
b. The MIND diet score, which ranges from 0 to 14, consists of 14 components, including green leafy vegetables, other vegetables, berries, whole grains, non-fried fish, non-fried poultry, beans, nuts, and olive oil.
